# Urinary Proteomic Signature of Mineralocorticoid Receptor Antagonism by Spironolactone: Evidence from the Randomized-Controlled HOMAGE and PRIORITY Trials

**DOI:** 10.1101/2023.04.05.23288107

**Authors:** Yu-Ling Yu, Viktor Rotbain-Curovic, Justyna Siwy, De-Wei An, Nete Tofte, Arantxa González, Morton K. Lindhardt, Tine W Hansen, Agnieszka Latosinska, João Pedro Ferreira, Pierpaolo Pellicori, Susana Ravassa, Beatrice Mariottoni, Job A.J. Verdonschot, Fozia Z. Ahmed, Johannes Petutschnigg, Patrick Rossignol, Stephane Heymans, Joe Cuthbert, Nicolas Girerd, Andrew L. Clark, Peter Verhamme, Tim S. Nawrot, Stefan Janssens, John G.F. Cleland, Faiez Zannad, Peter Rossing, Javier Díez, Harald Mischak, Jan A. Staessen HOMAGE and PRIORITY Investigators

## Abstract

**BACKGROUND:** Mineralocorticoid receptor (MR) activation induces fibrosis. Urinary proteomic profiling (UPP) detects thousands of sequenced peptides, mainly derived from collagen. No previous study applied UPP to generate insights in the antifibrotic actions of MR antagonism.

**METHODS:** Based on urine sample availability, subsets of the open HOMAGE trial (n=290; 23.8% women; median age: 73 years) and the double-blind PRIORITY trial (n=110; 21.8% women; 64 years) were analyzed as discovery and replication data sources. In the open HOMAGE trial, patients at risk of heart failure were randomized to usual therapy or usual therapy combined with spironolactone 25-50 mg/d. In the double-blind PRIORITY trial, type-2 diabetic patients with normal renal function were randomized to placebo or spironolactone 25 mg/d, both given on top of usual therapy. UPP relied on capillary electrophoresis coupled with mass spectrometry. In HOMAGE, the PICP/CITP ratio was calculated from serum PICP and serum CITP, which are markers of type-1 collagen synthesis and degradation, respectively. After rank-normalization of the biomarker distributions, between-group differences in the biomarker changes were analyzed by multivariable models. Correlations between the changes in urinary peptides and in serum CITP, derived from mature type-1 collagen, were compared between groups, using Fisher Z transform.

**RESULTS:** In the HOMAGE and PRIORITY analytical subsets, patients had detectable signals of 1498 urinary peptides. Follow-up totaled 9 months in HOMAGE and was 30 months (median) in PRIORITY. All changes in urinary peptides that remained significantly different (*P*<0.05) between randomization groups after accounting for baseline levels, covariables and multiple testing were collagen fragments. In HOMAGE and PRIORITY spironolactone reduced 16/27 and 10/13 collagen-derived urinary peptides. In HOMAGE, from baseline to 9 months, serum PICP and PICP/CITP decreased from 79.0 to 75.4 μg/L and from 21.3 to 18.3, respectively (*P*≤0.0129). Correlations between changes from baseline to follow-up in urinary type-1 collagen fragments and CITP were positive often reaching significance if fragments increased during follow-up, but were nonsignificant if fragments decreased during follow-up. There were no between-group differences in these correlations.

**CONCLUSIONS:** MR antagonism predominantly reduces collagen-derived urinary peptides. Inhibition of collagen synthesis by lowering the amount available for breakdown may be a contributing mechanism.

**Clinical Perspective:** *What Is New?:*

- Few studies addressed the association between urinary and serum markers of fibrosis and how MR antagonism influences urinary peptides derived from collagen.
- MR antagonism reduces collagen-derived urinary peptides. Inhibition of type-1 collagen synthesis by lowering the amount available for breakdown may be a contributing mechanism.
- Correlations between changes from baseline to follow-up in type-1 collagen and in CITP were positive if fragments increased during follow-up and nonsignificant if fragments decreased.

*What Are the Clinical Implications?:*

- Spironolactone inhibits fibrosis, supporting the use of MRAs in patients at risk of heart failure or chronic kidney disease.
- UPP profiling opens new research perspectives in documenting the antifibrotic properties of novel drug classes, such as nonsteroidal MR antagonists or sodium-glucose cotransporter-2 inhibitors.
- The development of novel medicines that would promote collagen degradation in addition to MRAs would strengthen the therapeutic armamentarium to modify fibrosis.

**GRAPHICAL ABSTRACT:** 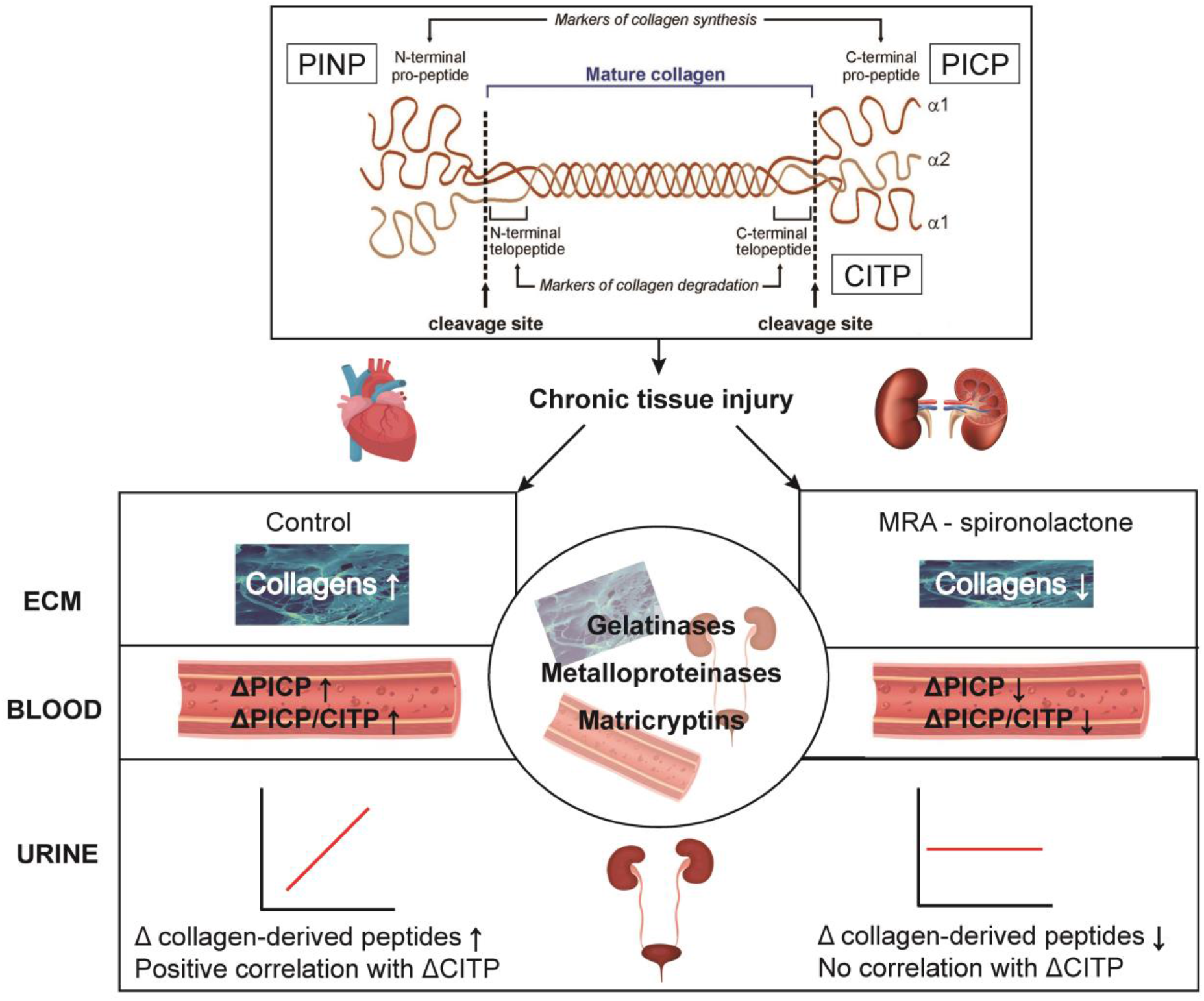

## INTRODUCTION

Fibrosis is the common response to inflammation and chronic tissue injury, such as occur with aging, hypertension, diabetes mellitus, or ischemia. Activation of the mineralocorticoid receptor (MR) by its ligands, aldosterone and cortisol, and non-ligands, such as hyperglycemia or salt loading, initiates a cascade of molecular events leading to cell growth, inappropriate expansion and disorganization of the extracellular matrix (ECM),^1^ which are common features shared by heart failure (HF) and chronic kidney disease (CKD).^2^

The Heart “Omics” in Ageing Study (HOMAGE) was an open-label randomized clinical trial with blinded endpoint evaluation in 527 patients at high HF risk.^3^ Patients were randomized to spironolactone 25-50 mg/d on top of usual therapy or usual therapy alone and were followed up for a maximum of 9 months.^3^ The objective was to test the effects of spironolactone on fibrosis markers to develop strategies to prevent HF. In patients on spironolactone, follow-up serum PICP and plasma NT-proBNP levels were lower, serum CITP levels higher, and the PINP-to-CITP ratio lower. These between-group differences were fairly constant up to the end of the trial for CITP (interaction *P*=0.065), whereas during follow-up serum PICP and the PICP-to-CITP ratio continued to decrease on spironolactone (*P*≤0.001). PICP and CITP are serum markers of collagen type-1 synthesis and degradation, respectively (**Figure S1**).^4^ The conclusion offered was that spironolactone may influence type-I collagen metabolism.^3^

Urine contains over 20,000 endogenous peptides, of which over 5000 have been sequenced, thereby identifying the parental proteins.^5^ The urinary proteomic profile (UPP) provides a whole-body signature of protein turnover.^6^ In uncontrolled observational studies, a decrease in urinary peptide fragments derived from type-1 collagen characterizes fibrosis.^7-9^ Given the renewed interest in MRAs,^10,11^ the current study investigated whether in a subset of the HOMAGE study population, UPP analysis provided novel information on the influence of MRA by spironolactone on the turnover of collagen I. To seek confirmation of the HOMAGE UPP findings, the UPP of patients at high risk of diabetic nephropathy enrolled in the double-blind placebo-controlled PRIORITY trial^12^ was also analyzed.

## METHODS

### Study Participants

HOMAGE was a multicenter open-label trial with blinded endpoint evaluation (registration number: NCT02556450).^3^ The HOMAGE trial was in 9 centers in the United Kingdom, France, Italy, Ireland, Germany and the Netherlands. Patients were screened in primary and secondary care. Each center had its own recruitment strategies under review of local ethics committees (see statistical analysis plan available at www.clinicaltrials.gov). Patients of either sex, aged ≥65 years (amended to ≥60 years) were eligible, provided that they were at increased risk of HF, because they had or were at high risk of coronary heart disease. Additionally, eligible patients had to have a serum NTproBNP of 125-1000 ng/L or a serum BNP of 35-280 ng/L. These ranges excluded patients at low HF risk as well as those with advanced disease requiring further investigation and treatment. Of 877 screened patients, 527 were randomized to spironolactone 25-50 mg per day (n=265) on top of usual treatment or usual treatment alone (n=262). The main analyses included 251 actively treated and 255 control patients. In the current analytical HOMAGE trial subset, urine samples were collected at baseline and at the 1-month and 9-month visits. Availability of urine samples and the detection of 1498 sequenced peptides in ≥30% of participants were the only selection criteria. Follow-up of all patients in the HOMAGE trial subset lasted 9 months. A flow chart depicting the derivation of the HOMAGE trial subset is available in the online only Data Supplement (**Figure S2**).

PRIORITY was a multicenter, prospective observational study with an embedded double-blind placebo-controlled trial (registration numbers: EudraCT 20120-004523-4 and NCT02040441).^12^ The PRIORITY study was conducted at 15 specialist centers in 10 European countries (Belgium, Czech Republic, Denmark, Germany, Greece, Italy, the Netherlands, North Macedonia, Spain, and the UK; a lists of sites and the local ethics approval numbers are available in reference 12. Eligible patients of either sex, aged 18-75 years, had type-2 diabetes with normal renal function and normoalbuminuria. At enrolment, all patients underwent a measurement of the multidimensional urinary proteomic classifier CKD273, which is predictive of progression of renal dysfunction.^13^ Those with a high-risk CKD273 score (>0.154) were randomized to spironolactone 25 mg per day (n=102) or matching placebo (n=107) on top of usual treatment.^12^ Median follow-up was 2.5 years (IQR: 2.0-3.0 years). As in HOMAGE, availability of urine samples and the detection of 1498 sequenced peptides in ≥30% of participants were the only selection criteria. Figure S3 shows the flow chart for the derivation of the analytical PRIORITY trial subset.

### Urinary and Circulating Biomarkers

Mosaiques-Diagnostics GmbH, Hannover, Germany (MOS) did the UPP profiling for all patients in both trials. The methods for sample preparation, CE-MS, peptide sequencing, and for the evaluation, calibration and quality control of the mass spectrometric data have been published^14^ and are described in detail in the online only Data Supplement (pp 2-4). In the CE-MS step, reference signals of 29 abundant endogenous urinary peptides were used as internal standards for calibration of signal intensity. This procedure is highly reproducible and addresses both analytical and dilution variances, such as the variability in renal function, in a single calibration step.^15^ A total of 1498 sequenced urinary peptides with a detectable signal in ≥30% of participants were analyzed. Undetectable peptides were set at the distribution minimum.^16^ eGFR was estimated from serum creatinine by the Chronic Kidney Disease Epidemiology Collaboration (CKD-EPI) formula.^17^ In HOMAGE participants, using methods described in the Data Supplement (p 5), serum was analyzed for PICP, a marker of type-1 collagen synthesis and CITP a marker of type-1 collagen degradation (**Figure S1**).^18^

### Statistical Analysis

For database management and statistical analysis, SAS software, version 9.4, maintenance level 5 was used (SAS Institute Inc., Cary, NC). Deviation from the normal distribution was assessed by the Shapiro-Wilk statistic. The distributions of serum biomarkers (**Figure S4**) and urinary peptides (**Figure S5**) were rank normalized, by sorting measurements from the smallest to the highest value and then applying the inverse cumulative normal function.^19^ Rank normalized variables have mean 0 and SD 1. For non-transformed and for rank normalized variables, the central tendency (spread) of the data were given as the median (IQR) or as the arithmetic mean (SD), respectively. Means were compared using the large-sample Z-test and proportions by the χ^2^ statistic or the Fisher exact test, as appropriate based on cell frequencies. Statistical tests were two-sided.

The UPP analysis was done according to predefined steps.^6^ Between-group differences in the biomarkers (spironolactone *vs* placebo) were examined, using general linear models with adjustment for the baseline level if the test involved follow-up data. Next the general linear models were multivariable adjusted. The covariables considered in HOMAGE were sex, age, BMI, and eGFR, smoking and drinking alcohol at enrolment, history of ischemic heart disease, and treatment at baseline and subsequent treatment changes during follow-up with antihypertensive, lipid-lowering, antiplatelet and antidiabetic drugs. The antihypertensive drugs considered as covariables were thiazide, thiazide-like, and loop diuretics, β-blockers, vasodilators (calcium-channel blockers and α-blockers), and inhibitors of the renin-angiotensin system (angiotensin-converting enzyme inhibitors and angiotensin receptor blockers). Lipid-lowering drugs included statins, fibrates and ezitimibe, and antiplatelet drugs aspirin and adenosine diphosphate receptor inhibitors. The antidiabetic drugs coded included insulin, metformin, sulfonylurea, dipeptyl peptidase-4 inhibitors, glucagon-like peptide-1 receptor antagonists, thiazolididiones and sodium-glucose cotransporter 2 inhibitors. In PRIORITY analyses, the available covariables were sex, age, BMI, eGFR, smoking, the duration of diabetes, history of cardiovascular disease, and treatment at baseline with antihypertensive or antidiabetic drugs.

The urinary peptide fragments to be retained in the analyses were selected in two steps. The first step involved selecting the urinary peptides with a different abundance at last follow-up among control and spironolactone-treated patients with a 2-sided significance of 0.01. Next, peptides keeping Benjamini-Hochberg-adjusted significance^20^ of <0.05 were carried through to further analysis. In the HOMAGE trial subset, Pearson correlation coefficients of the changes from baseline to follow-up in the urinary peptides regressed on the serum biomarkers were compared between treatment groups (control *vs* spironolactone), using Fisher Z-transformation.^21^ Between-group comparisons of the slopes of these associations were tested by linear regression models, including randomization group, the changes in the serum biomarker, and the interaction term between randomization group and the changes in the serum biomarker.^22^ These analyses were confined to CITP, because the peptide fragments detectable in urine are derived from mature type-1 collagen.^9^

## RESULTS

### Patient Characteristics

The 290 patients in the HOMAGE trial subset included 69 (23.8%) women. At baseline, median values (IQR) were 73 (69-78) years for age, 28 (25-31) kg/m^2^ for BMI, 136 (125-152) mm Hg for systolic BP, 76 (70-84) for diastolic BP, and 67 (58-79) mL/min/1.73 m^2^ for eGFR. The number of participants with hypertension, diabetes or ischemic heart disease amounted to 217 (74.8%), 64 (22.1%), and 235 (81.0%), respectively. The HOMAGE patients were intensively treated with antihypertensive agents (n=280 [96.6%]) and lipid-lowering drugs (n=257 [88.6%], mainly statins (n=251 [86.6%]), antiplatelet drugs (n=218 [75.2%]), and antidiabetic agents (n=105 [36.2%]), insulin in 10 (3.4%) cases. The analytical subset of HOMAGE patients, randomized to control (n=144) or spironolactone (n=146), were well balanced with regard to risk factors, clinical characteristics, routine biochemistry, and serum PICP, CITP and the PICP/CITP ratio (**Table 1**). Compared with the 215 HOMAGE patients not included in the present analyses, the 290 patients analyzed had a higher risk profile (**Table S1**).

**Table 1.**
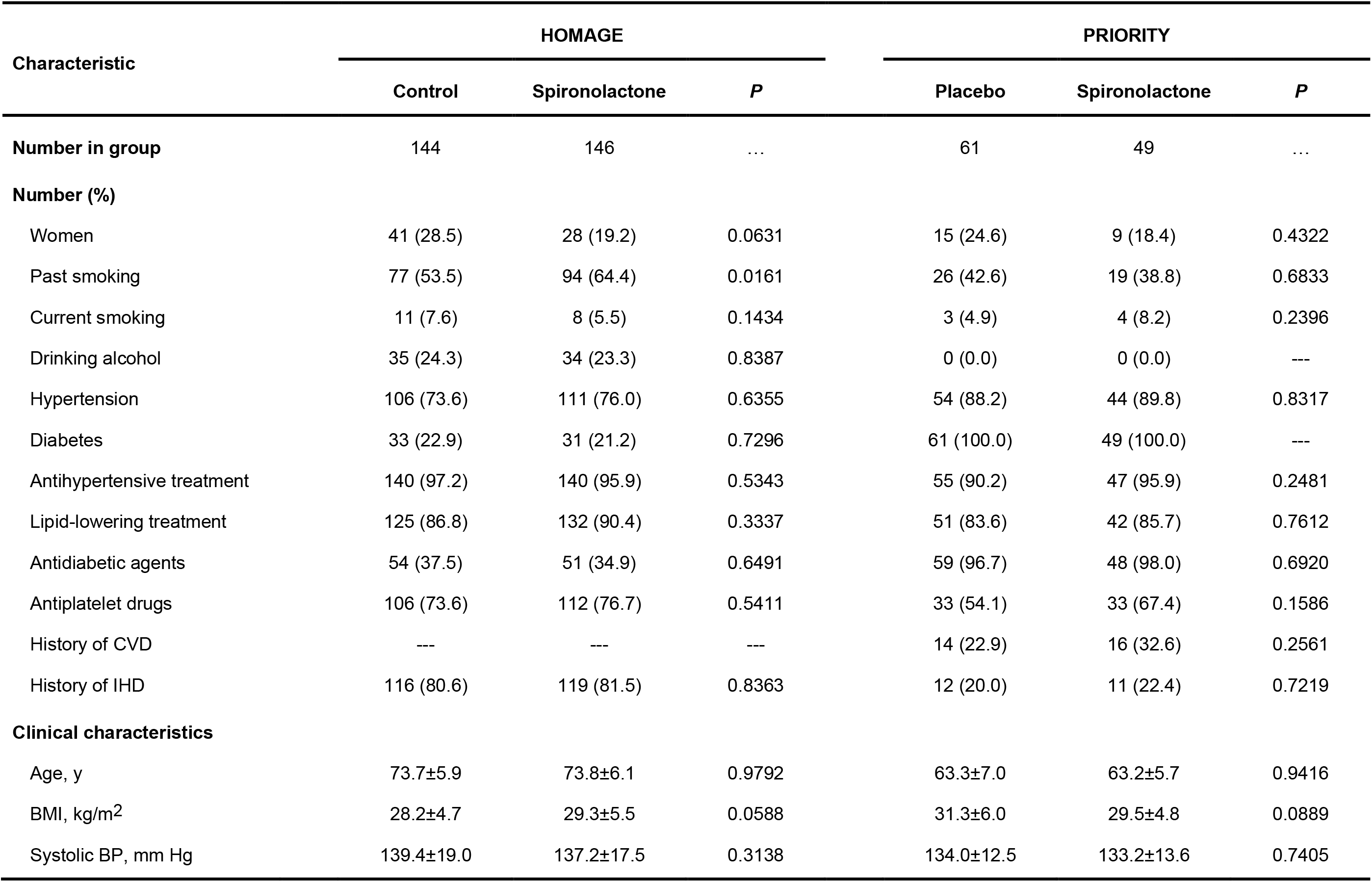
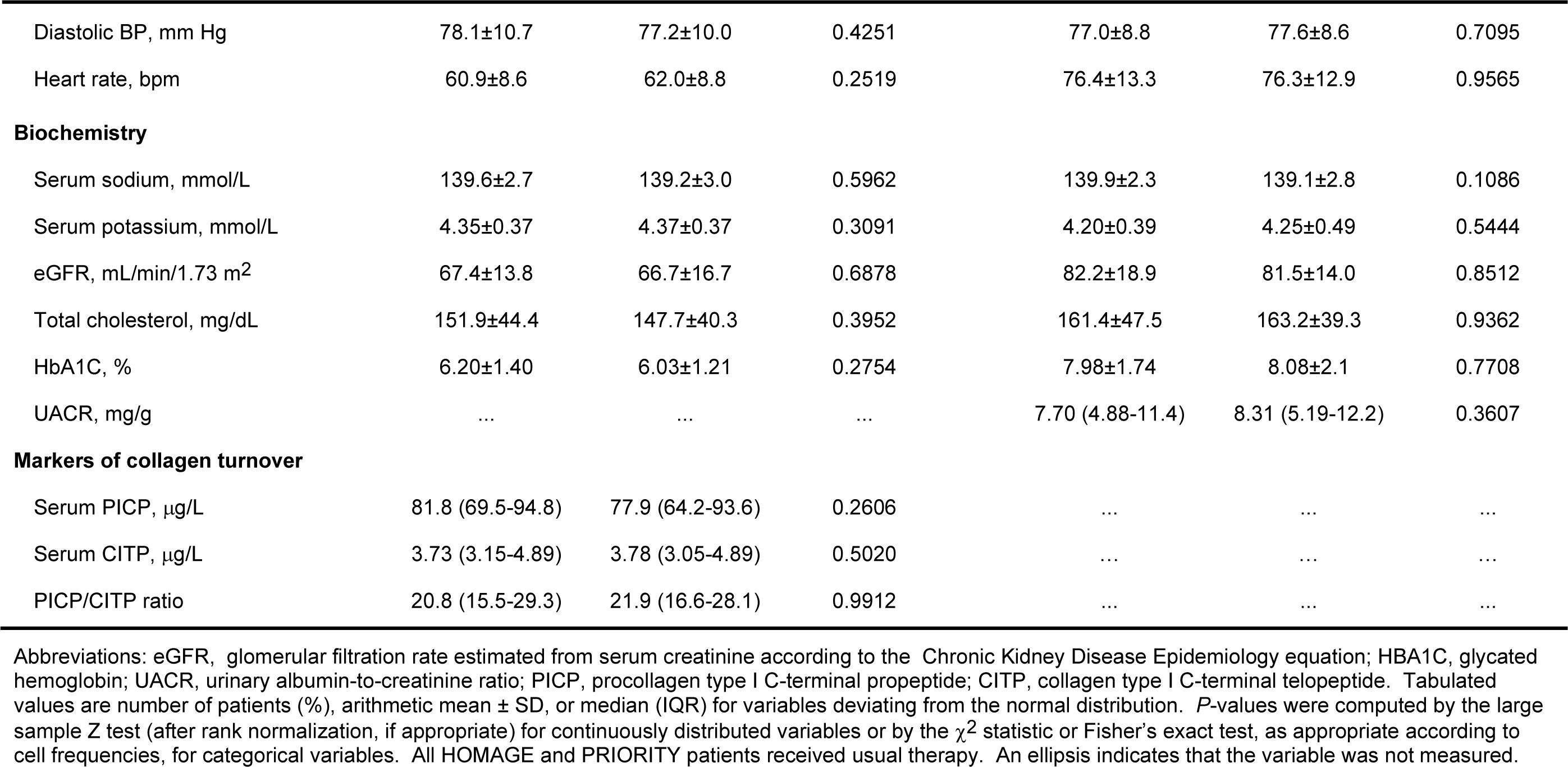
Baseline Characteristics of Patients.

| Characteristic | HOMAGE |  |  | PRIORITY |  |  |
| --- | --- | --- | --- | --- | --- | --- |
|  | Control | Spironolactone | *P* | Placebo | Spironolactone | *P* |
| **Number in group** | 144 | 146 | ... | 61 | 49 | ... |
| **Number (%)** |  |  |  |  |  |  |
| Women | 41 (28.5) | 28 (19.2) | 0.0631 | 15 (24.6) | 9 (18.4) | 0.4322 |
| Past smoking | 77 (53.5) | 94 (64.4) | 0.0161 | 26 (42.6) | 19 (38.8) | 0.6833 |
| Current smoking | 11 (7.6) | 8 (5.5) | 0.1434 | 3 (4.9) | 4 (8.2) | 0.2396 |
| Drinking alcohol | 35 (24.3) | 34 (23.3) | 0.8387 | 0 (0.0) | 0 (0.0) | --- |
| Hypertension | 106 (73.6) | 111 (76.0) | 0.6355 | 54 (88.2) | 44 (89.8) | 0.8317 |
| Diabetes | 33 (22.9) | 31 (21.2) | 0.7296 | 61 (100.0) | 49 (100.0) | --- |
| Antihypertensive treatment | 140 (97.2) | 140 (95.9) | 0.5343 | 55 (90.2) | 47 (95.9) | 0.2481 |
| Lipid-lowering treatment | 125 (86.8) | 132 (90.4) | 0.3337 | 51 (83.6) | 42 (85.7) | 0.7612 |
| Antidiabetic agents | 54 (37.5) | 51 (34.9) | 0.6491 | 59 (96.7) | 48 (98.0) | 0.6920 |
| Antiplatelet drugs | 106 (73.6) | 112 (76.7) | 0.5411 | 33 (54.1) | 33 (67.4) | 0.1586 |
| History of CVD | --- | --- | --- | 14 (22.9) | 16 (32.6) | 0.2561 |
| History of IHD | 116 (80.6) | 119 (81.5) | 0.8363 | 12 (20.0) | 11 (22.4) | 0.7219 |
| **Clinical characteristics** |  |  |  |  |  |  |
| Age, y | 73.7±5.9 | 73.8±6.1 | 0.9792 | 63.3±7.0 | 63.2±5.7 | 0.9416 |
| BMI, kg/m<sup>2</sup> | 28.2±4.7 | 29.3±5.5 | 0.0588 | 31.3±6.0 | 29.5±4.8 | 0.0889 |
| Systolic BP, mm Hg | 139.4±19.0 | 137.2±17.5 | 0.3138 | 134.0±12.5 | 133.2±13.6 | 0.7405 |

**Table 1 Baseline Characteristics of Patients (Continued)**
| Characteristic | HOMAGE |  |  | PRIORITY |  |  |
| --- | --- | --- | --- | --- | --- | --- |
|  | Control | Spironolactone | *P* | Placebo | Spironolactone | *P* |
| Diastolic BP, mm Hg | 78.1±10.7 | 77.2±10.0 | 0.4251 | 77.0±8.8 | 77.6±8.6 | 0.7095 |
| Heart rate, bpm | 60.9±8.6 | 62.0±8.8 | 0.2519 | 76.4±13.3 | 76.3±12.9 | 0.9565 |
| **Biochemistry** |  |  |  |  |  |  |
| Serum sodium, mmol/L | 139.6±2.7 | 139.2±3.0 | 0.5962 | 139.9±2.3 | 139.1±2.8 | 0.1086 |
| Serum potassium, mmol/L | 4.35±0.37 | 4.37±0.37 | 0.3091 | 4.20±0.39 | 4.25±0.49 | 0.5444 |
| eGFR, mL/min/1.73 m<sup>2</sup> | 67.4±13.8 | 66.7±16.7 | 0.6878 | 82.2±18.9 | 81.5±14.0 | 0.8512 |
| Total cholesterol, mg/dL | 151.9±44.4 | 147.7±40.3 | 0.3952 | 161.4±47.5 | 163.2±39.3 | 0.9362 |
| HbA1C, % | 6.20±1.40 | 6.03±1.21 | 0.2754 | 7.98±1.74 | 8.08±2.1 | 0.7708 |
| UACR, mg/g | ... | ... | ... | 7.70 (4.88-11.4) | 8.31 (5.19-12.2) | 0.3607 |
| **Markers of collagen turnover** |  |  |  |  |  |  |
| Serum PICP, µg/L | 81.8 (69.5-94.8) | 77.9 (64.2-93.6) | 0.2606 | ... | ... | ... |
| Serum CITP, µg/L | 3.73 (3.15-4.89) | 3.78 (3.05-4.89) | 0.5020 | ... | ... | ... |
| PICP/CITP ratio | 20.8 (15.5-29.3) | 21.9 (16.6-28.1) | 0.9912 | ... | ... | ... |

Abbreviations: eGFR, glomerular filtration rate estimated from serum creatinine according to the Chronic Kidney Disease Epidemiology equation; HbA1C, glycated hemoglobin; UACR, urinary albumin-to-creatinine ratio; PICP, procollagen type I C-terminal propeptide; CITP, collagen type I C-terminal telopeptide. Tabulated values are number of patients (%), arithmetic mean ± SD, or median (IQR) for variables deviating from the normal distribution. *P*-values were computed by the large sample Z test (after rank normalization, if appropriate) for continuously distributed variables or by the $\chi^2$ statistic or Fisher's exact test, as appropriate according to cell frequencies, for categorical variables. All HOMAGE and PRIORITY patients received usual therapy. An ellipsis indicates that the variable was not measured.

The 110 PRIORITY participants available for analysis included 24 (21.8%) women. At baseline, median values (IQR) were 64.0 (59.0-68.0) years for age, 29.6 (26.9-33.3) kg/m^2^ for BMI, 132.0 (125.0-144.0) mm Hg for systolic BP, 78.5 (71.0-83.0) mm Hg for diastolic BP, and 84.0 (71.6-95.7) mL/min/1.73 m^2^ for eGFR. All patients were normoalbuminuric (median urinary albumin-to-creatinine ratio: 8.29 mg/g [IQR: 5.03-12.2]). The number of participants with hypertension or ischemic heart disease amounted to 98 (89.1%) and 23 (20.9%), respectively. The PRIORITY participants were treated with antihypertensiva agents (n=102 [97.3%]), lipid-lowering drugs (n=93 [84.5%], mainly statins (n=91 [82.7%]), antiplatelet drugs (n=66 [60.0%]), aspirin in most cases (n=63 [57.3%]), and antidiabetic agents (n=107 [97.3%]), insulin in 37 (33.6%) patients. The PRIORITY patients in the analytical subset, randomized to placebo (n=61) or spironolactone (n=49) had similar characteristics (**Table 1**). Compared with the 99 PRIORITY participants not included in the present analyses, the 110 analyzed patients had a higher risk profile (**Table S1**).

### Urinary Peptides

The 1498 sequenced urinary peptides identified 212 proteins. Peptides derived from albumin, β_2_-microglobulin and the fibrinogen α-chain were excluded from further analysis given their high concentration in the blood, uromodulin because of its renal origin, and osteopontin as prominent component of mineralized extracellular matrixes. Given the balanced characteristics of patients randomized to control or spironolactone, the between-group differences in the urinary and serum biomarkers and associated significance levels were largely similar irrespective of adjustment.

While accounting for multiple testing, adjusted models of the HOMAGE trial subset data demonstrated that 27 peptides had a different abundance at last follow-up in controls compared to patients randomized to spironolactone (**Table 2**). The proteins identified were all collagens: COL1A1 (11 peptides), COL2A1 (1), COL3A1 (4), COL4A1 (1), COL6A1 (1), COL7A1 (1), COL1A2 (2), COL4A2 (1), COL5A2 (1), COL11A2 (1), COL4A3 (1), COL5A3 (1), and COL4A6 (1). With adjustment for multiple testing, in the PRIORITY trial subset, 14 peptides showed a different abundance at last follow-up (**Table 2**): COL1A1 (2 fragments), COL2A1 (1), COL3A1 (1), COL5A1 (1), COL6A1 (1), COL11A1 (1), COL15A1 (1), COL16A1 (1), COL22A1 (1), COL1A2 (1), COL5A2 (1), COL4A4 (1), and COL4A6 (1). For each peptide listed in **Table 2**, the median of the ratio (spironolactone/control) of the change in peptide abundance from baseline to last follow-up is shown. Fragment e06373 (COL4A6) was significant in both trials, but had an opposite ratio in HOMAGE and PRIORITY: 0.87 *vs* 2.13, respectively. Of the other 38 peptide fragments, 23 (60.5%) showed concordant ratios in both trials (**Table 2**). **Table S2** lists the amino-acid sequence of the peptides retained in the HOMAGE and PRIORITY analyses and the protein from which they were derived. At baseline, the abundance of the selected urinary peptides was similar in both treatment groups in the HOMAGE and PRIORITY subsets (**Table S3**).

**Table 2.**
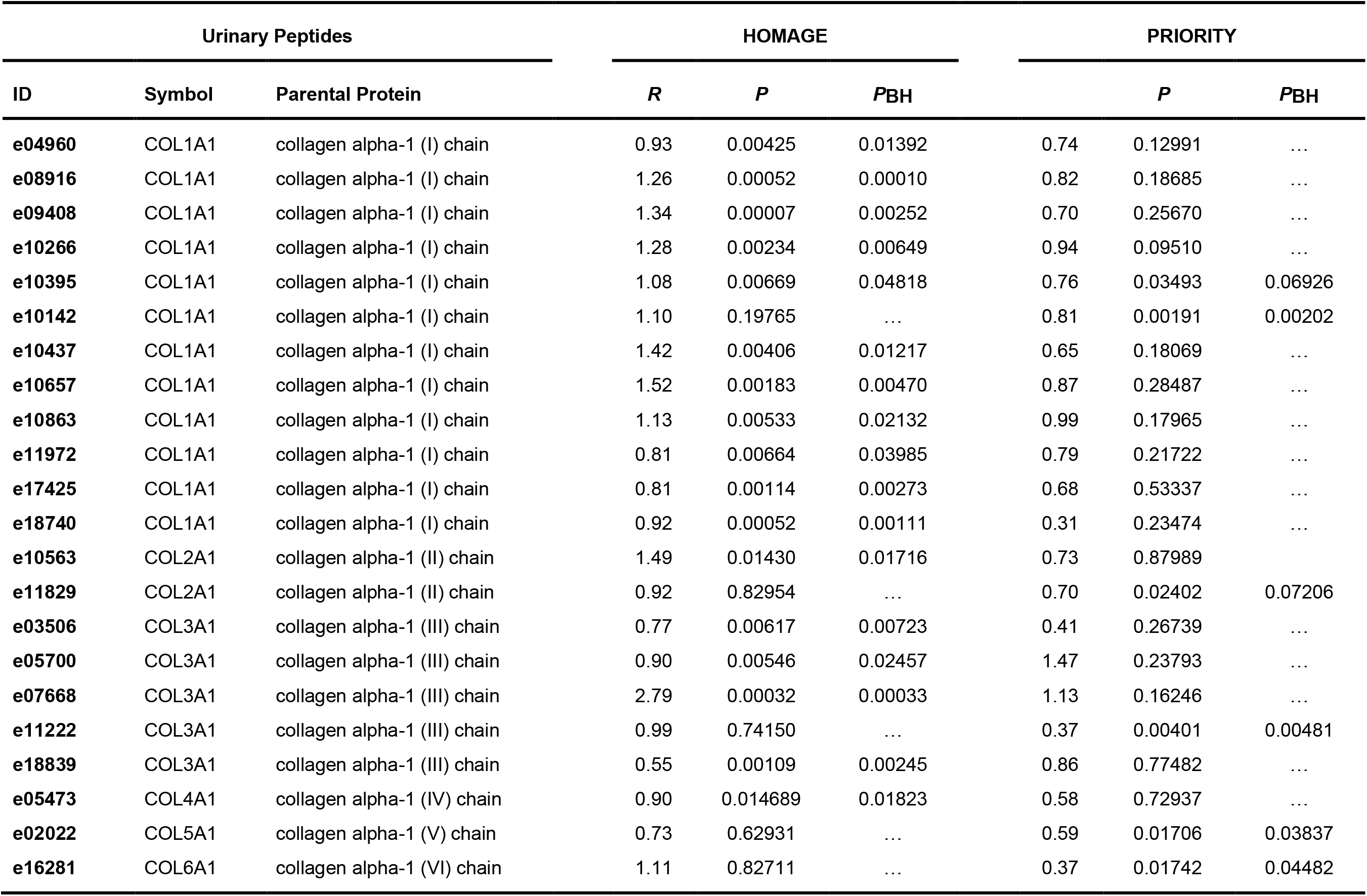
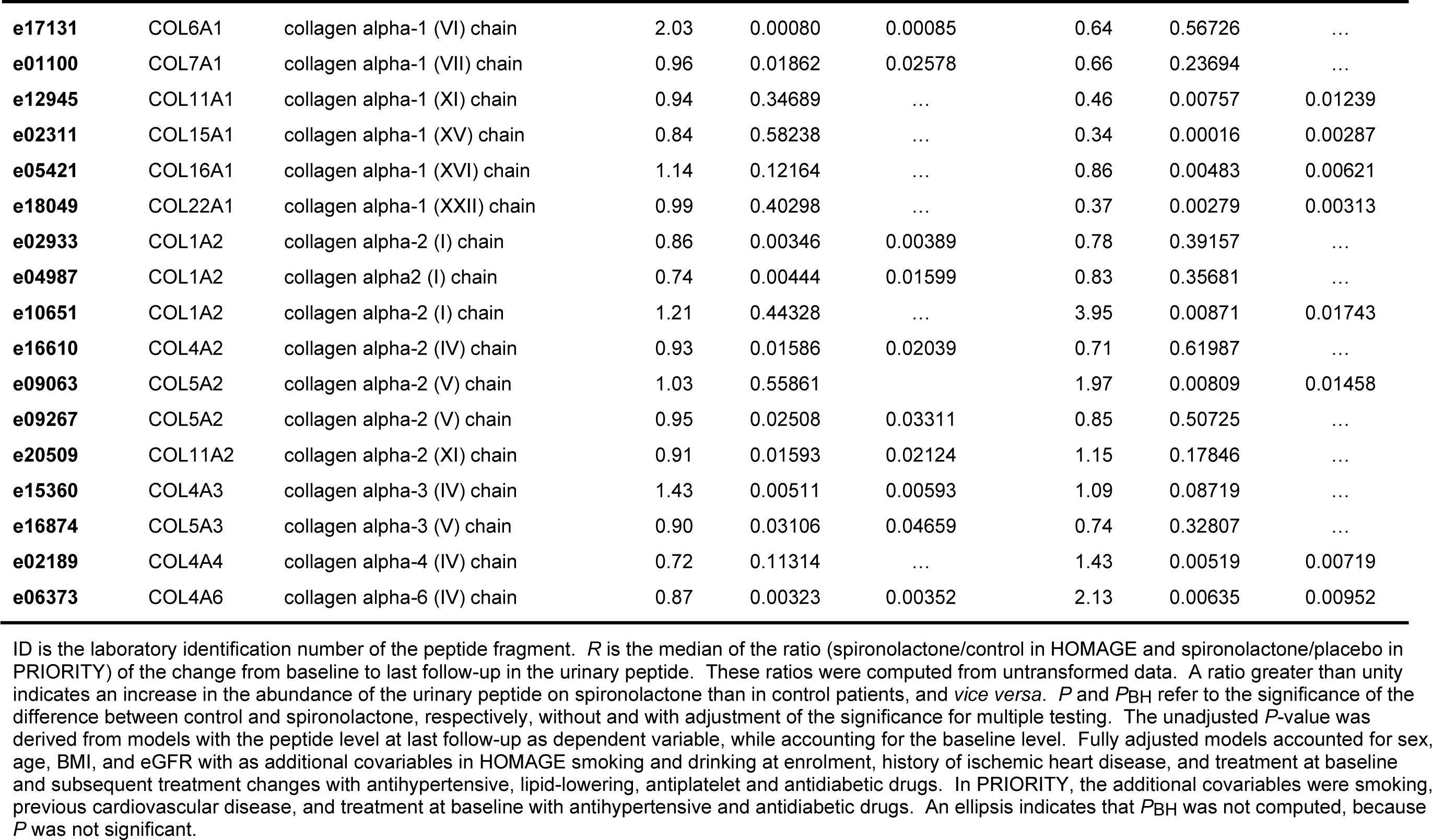
Urinary Peptides with Different Abundance on Spironolactone versus Control Retained in the Analyses.

| Urinary Peptides |  |  | HOMAGE |  |  | PRIORITY |  |  |
| --- | --- | --- | --- | --- | --- | --- | --- | --- |
| ID | Symbol | Parental Protein | *R* | *P* | *PBH* |  | *P* | *PBH* |
| e04960 | COL1A1 | collagen alpha-1 (I) chain | 0.93 | 0.00425 | 0.01392 | 0.74 | 0.12991 | ... |
| e08916 | COL1A1 | collagen alpha-1 (I) chain | 1.26 | 0.00052 | 0.00010 | 0.82 | 0.18685 | ... |
| e09408 | COL1A1 | collagen alpha-1 (I) chain | 1.34 | 0.00007 | 0.00252 | 0.70 | 0.25670 | ... |
| e10266 | COL1A1 | collagen alpha-1 (I) chain | 1.28 | 0.00234 | 0.00649 | 0.94 | 0.09510 | ... |
| e10395 | COL1A1 | collagen alpha-1 (I) chain | 1.08 | 0.00669 | 0.04818 | 0.76 | 0.03493 | 0.06926 |
| e10142 | COL1A1 | collagen alpha-1 (I) chain | 1.10 | 0.19765 | ... | 0.81 | 0.00191 | 0.00202 |
| e10437 | COL1A1 | collagen alpha-1 (I) chain | 1.42 | 0.00406 | 0.01217 | 0.65 | 0.18069 | ... |
| e10657 | COL1A1 | collagen alpha-1 (I) chain | 1.52 | 0.00183 | 0.00470 | 0.87 | 0.28487 | ... |
| e10863 | COL1A1 | collagen alpha-1 (I) chain | 1.13 | 0.00533 | 0.02132 | 0.99 | 0.17965 | ... |
| e11972 | COL1A1 | collagen alpha-1 (I) chain | 0.81 | 0.00664 | 0.03985 | 0.79 | 0.21722 | ... |
| e17425 | COL1A1 | collagen alpha-1 (I) chain | 0.81 | 0.00114 | 0.00273 | 0.68 | 0.53337 | ... |
| e18740 | COL1A1 | collagen alpha-1 (I) chain | 0.92 | 0.00052 | 0.00111 | 0.31 | 0.23474 | ... |
| e10563 | COL2A1 | collagen alpha-1 (II) chain | 1.49 | 0.01430 | 0.01716 | 0.73 | 0.87989 | ... |
| e11829 | COL2A1 | collagen alpha-1 (II) chain | 0.92 | 0.82954 | ... | 0.70 | 0.02402 | 0.07206 |
| e03506 | COL3A1 | collagen alpha-1 (III) chain | 0.77 | 0.00617 | 0.00723 | 0.41 | 0.26739 | ... |
| e05700 | COL3A1 | collagen alpha-1 (III) chain | 0.90 | 0.00546 | 0.02457 | 1.47 | 0.23793 | ... |
| e07668 | COL3A1 | collagen alpha-1 (III) chain | 2.79 | 0.00032 | 0.00033 | 1.13 | 0.16246 | ... |
| e11222 | COL3A1 | collagen alpha-1 (III) chain | 0.99 | 0.74150 | ... | 0.37 | 0.00401 | 0.00481 |
| e18839 | COL3A1 | collagen alpha-1 (III) chain | 0.55 | 0.00109 | 0.00245 | 0.86 | 0.77482 | ... |
| e05473 | COL4A1 | collagen alpha-1 (IV) chain | 0.90 | 0.014689 | 0.01823 | 0.58 | 0.72937 | ... |
| e02022 | COL5A1 | collagen alpha-1 (V) chain | 0.73 | 0.62931 | ... | 0.59 | 0.01706 | 0.03837 |
| e16281 | COL6A1 | collagen alpha-1 (VI) chain | 1.11 | 0.82711 | ... | 0.37 | 0.01742 | 0.04482 |

**Table 2 Urinary Peptides with Different Abundance on Spironolactone versus Placebo Retained in the Analyses (Continued)**
| Urinary Peptides |  |  | HOMAGE |  |  | PRIORITY |  |  |
| --- | --- | --- | --- | --- | --- | --- | --- | --- |
| ID | Symbol | Parental Protein | *R* | *P* | *P<sub>BH</sub>* | *R* | *P* | *P<sub>BH</sub>* |
| **e17131** | COL6A1 | collagen alpha-1 (VI) chain | 2.03 | 0.00080 | 0.00085 | 0.64 | 0.56726 | ... |
| **e01100** | COL7A1 | collagen alpha-1 (VII) chain | 0.96 | 0.01862 | 0.02578 | 0.66 | 0.23694 | ... |
| **e12945** | COL11A1 | collagen alpha-1 (XI) chain | 0.94 | 0.34689 | ... | 0.46 | 0.00757 | 0.01239 |
| **e02311** | COL15A1 | collagen alpha-1 (XV) chain | 0.84 | 0.58238 | ... | 0.34 | 0.00016 | 0.00287 |
| **e05421** | COL16A1 | collagen alpha-1 (XVI) chain | 1.14 | 0.12164 | ... | 0.86 | 0.00483 | 0.00621 |
| **e18049** | COL22A1 | collagen alpha-1 (XXII) chain | 0.99 | 0.40298 | ... | 0.37 | 0.00279 | 0.00313 |
| **e02933** | COL1A2 | collagen alpha-2 (I) chain | 0.86 | 0.00346 | 0.00389 | 0.78 | 0.39157 | ... |
| **e04987** | COL1A2 | collagen alpha2 (I) chain | 0.74 | 0.00444 | 0.01599 | 0.83 | 0.35681 | ... |
| **e10651** | COL1A2 | collagen alpha-2 (I) chain | 1.21 | 0.44328 | ... | 3.95 | 0.00871 | 0.01743 |
| **e16610** | COL4A2 | collagen alpha-2 (IV) chain | 0.93 | 0.01586 | 0.02039 | 0.71 | 0.61987 | ... |
| **e09063** | COL5A2 | collagen alpha-2 (V) chain | 1.03 | 0.55861 | ... | 1.97 | 0.00809 | 0.01458 |
| **e09267** | COL5A2 | collagen alpha-2 (V) chain | 0.95 | 0.02508 | 0.03311 | 0.85 | 0.50725 | ... |
| **e20509** | COL11A2 | collagen alpha-2 (XI) chain | 0.91 | 0.01593 | 0.02124 | 1.15 | 0.17846 | ... |
| **e15360** | COL4A3 | collagen alpha-3 (IV) chain | 1.43 | 0.00511 | 0.00593 | 1.09 | 0.08719 | ... |
| **e16874** | COL5A3 | collagen alpha-3 (V) chain | 0.90 | 0.03106 | 0.04659 | 0.74 | 0.32807 | ... |
| **e02189** | COL4A4 | collagen alpha-4 (IV) chain | 0.72 | 0.11314 | ... | 1.43 | 0.00519 | 0.00719 |
| **e06373** | COL4A6 | collagen alpha-6 (IV) chain | 0.87 | 0.00323 | 0.00352 | 2.13 | 0.00635 | 0.00952 |

ID is the laboratory identification number of the peptide fragment. *R* is the median of the ratio (spironolactone/control in HOMAGE and spironolactone/placebo in PRIORITY) of the change from baseline to last follow-up in the urinary peptide. These ratios were computed from untransformed data. A ratio greater than unity indicates an increase in the abundance of the urinary peptide on spironolactone than in control patients, and *vice versa*. *P* and *P<sub>BH</sub>* refer to the significance of the difference between control and spironolactone, respectively, without and with adjustment of the significance for multiple testing. The unadjusted *P*-value was derived from models with the peptide level at last follow-up as dependent variable, while accounting for the baseline level. Fully adjusted models accounted for sex, age, BMI, and eGFR with as additional covariables in HOMAGE smoking and drinking at enrolment, history of ischemic heart disease, and treatment at baseline and subsequent treatment changes with antihypertensive, lipid-lowering, antiplatelet and antidiabetic drugs. In PRIORITY, the additional covariables were smoking, previous cardiovascular disease, and treatment at baseline with antihypertensive and antidiabetic drugs. An ellipsis indicates that *P<sub>BH</sub>* was not computed, because *P* was not significant.

**Table 3.**
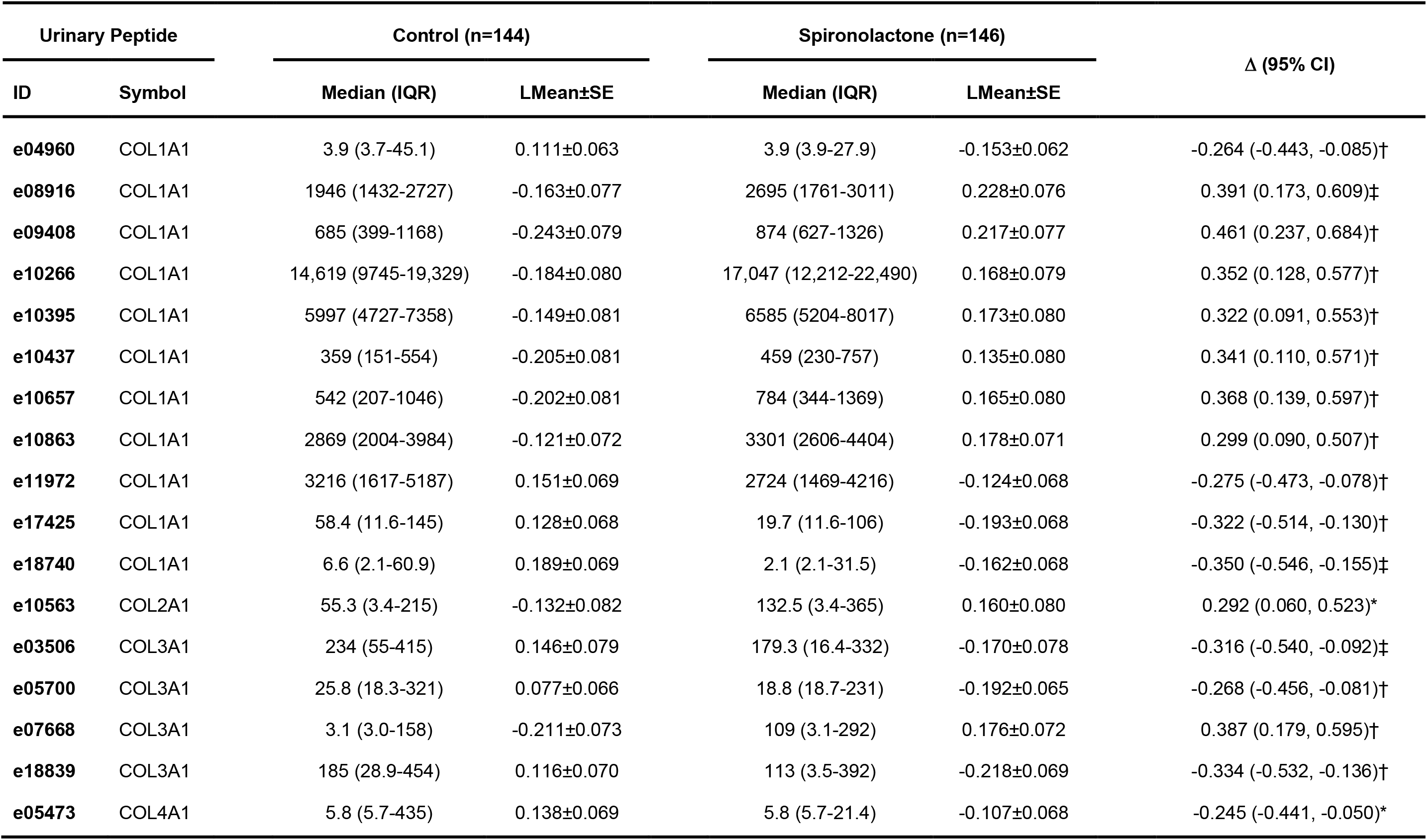
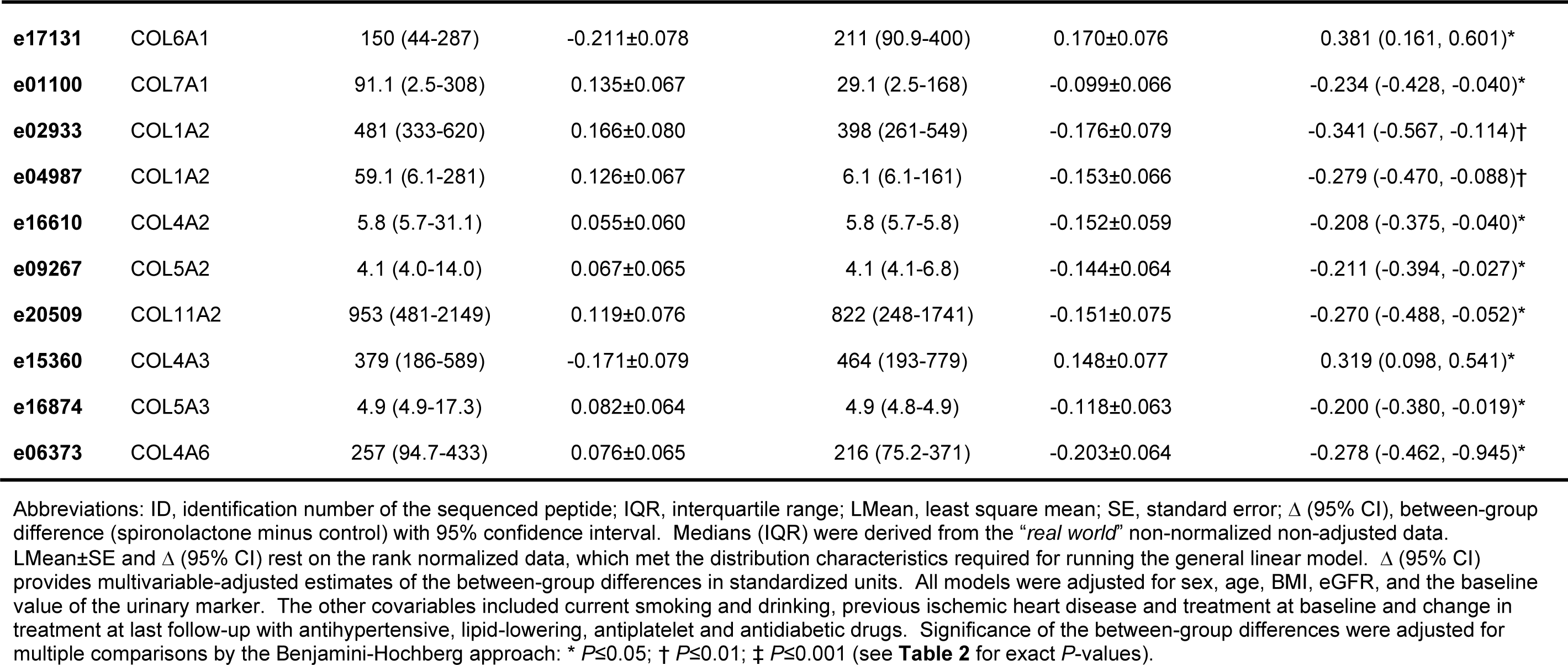
Between-Group Differences in the Urinary Peptide Levels at Last Follow-Up in the HOMAGE Trial Subset.

| Urinary Peptide | | Control (n=144) | | Spironolactone (n=146) | | $\Delta$ (95% CI) |
| --- | --- | --- | --- | --- | --- | --- |
| ID | Symbol | Median (IQR) | LMean±SE | Median (IQR) | LMean±SE | |
| e04960 | COL1A1 | 3.9 (3.7-45.1) | 0.111±0.063 | 3.9 (3.9-27.9) | -0.153±0.062 | -0.264 (-0.443, -0.085)† |
| e08916 | COL1A1 | 1946 (1432-2727) | -0.163±0.077 | 2695 (1761-3011) | 0.228±0.076 | 0.391 (0.173, 0.609)‡ |
| e09408 | COL1A1 | 685 (399-1168) | -0.243±0.079 | 874 (627-1326) | 0.217±0.077 | 0.461 (0.237, 0.684)† |
| e10266 | COL1A1 | 14,619 (9745-19,329) | -0.184±0.080 | 17,047 (12,212-22,490) | 0.168±0.079 | 0.352 (0.128, 0.577)† |
| e10395 | COL1A1 | 5997 (4727-7358) | -0.149±0.081 | 6585 (5204-8017) | 0.173±0.080 | 0.322 (0.091, 0.553)† |
| e10437 | COL1A1 | 359 (151-554) | -0.205±0.081 | 459 (230-757) | 0.135±0.080 | 0.341 (0.110, 0.571)† |
| e10657 | COL1A1 | 542 (207-1046) | -0.202±0.081 | 784 (344-1369) | 0.165±0.080 | 0.368 (0.139, 0.597)† |
| e10863 | COL1A1 | 2869 (2004-3984) | -0.121±0.072 | 3301 (2606-4404) | 0.178±0.071 | 0.299 (0.090, 0.507)† |
| e11972 | COL1A1 | 3216 (1617-5187) | 0.151±0.069 | 2724 (1469-4216) | -0.124±0.068 | -0.275 (-0.473, -0.078)† |
| e17425 | COL1A1 | 58.4 (11.6-145) | 0.128±0.068 | 19.7 (11.6-106) | -0.193±0.068 | -0.322 (-0.514, -0.130)† |
| e18740 | COL1A1 | 6.6 (2.1-60.9) | 0.189±0.069 | 2.1 (2.1-31.5) | -0.162±0.068 | -0.350 (-0.546, -0.155)‡ |
| e10563 | COL2A1 | 55.3 (3.4-215) | -0.132±0.082 | 132.5 (3.4-365) | 0.160±0.080 | 0.292 (0.060, 0.523)* |
| e03506 | COL3A1 | 234 (55-415) | 0.146±0.079 | 179.3 (16.4-332) | -0.170±0.078 | -0.316 (-0.540, -0.092)‡ |
| e05700 | COL3A1 | 25.8 (18.3-321) | 0.077±0.066 | 18.8 (18.7-231) | -0.192±0.065 | -0.268 (-0.456, -0.081)† |
| e07668 | COL3A1 | 3.1 (3.0-158) | -0.211±0.073 | 109 (3.1-292) | 0.176±0.072 | 0.387 (0.179, 0.595)† |
| e18839 | COL3A1 | 185 (28.9-454) | 0.116±0.070 | 113 (3.5-392) | -0.218±0.069 | -0.334 (-0.532, -0.136)† |
| e05473 | COL4A1 | 5.8 (5.7-435) | 0.138±0.069 | 5.8 (5.7-21.4) | -0.107±0.068 | -0.245 (-0.441, -0.050)* |

**Table 3 Between-Group Differences in the Urinary Peptide Levels at Last Follow-Up in the HOMAGE Trial Subset (Continued)**
| Urinary Peptide | | Placebo (n=144) | | Spironolactone (n=146) | | $\Delta$ (95% CI) |
| --- | --- | --- | --- | --- | --- | --- |
| ID | Symbol | Median (IQR) | LMean±SE | Median (IQR) | LMean±SE | |
| e17131 | COL6A1 | 150 (44-287) | -0.211±0.078 | 211 (90.9-400) | 0.170±0.076 | 0.381 (0.161, 0.601)* |
| e01100 | COL7A1 | 91.1 (2.5-308) | 0.135±0.067 | 29.1 (2.5-168) | -0.099±0.066 | -0.234 (-0.428, -0.040)* |
| e02933 | COL1A2 | 481 (333-620) | 0.166±0.080 | 398 (261-549) | -0.176±0.079 | -0.341 (-0.567, -0.114)† |
| e04987 | COL1A2 | 59.1 (6.1-281) | 0.126±0.067 | 6.1 (6.1-161) | -0.153±0.066 | -0.279 (-0.470, -0.088)† |
| e16610 | COL4A2 | 5.8 (5.7-31.1) | 0.055±0.060 | 5.8 (5.7-5.8) | -0.152±0.059 | -0.208 (-0.375, -0.040)* |
| e09267 | COL5A2 | 4.1 (4.0-14.0) | 0.067±0.065 | 4.1 (4.1-6.8) | -0.144±0.064 | -0.211 (-0.394, -0.027)* |
| e20509 | COL11A2 | 953 (481-2149) | 0.119±0.076 | 822 (248-1741) | -0.151±0.075 | -0.270 (-0.488, -0.052)* |
| e15360 | COL4A3 | 379 (186-589) | -0.171±0.079 | 464 (193-779) | 0.148±0.077 | 0.319 (0.098, 0.541)* |
| e16874 | COL5A3 | 4.9 (4.9-17.3) | 0.082±0.064 | 4.9 (4.8-4.9) | -0.118±0.063 | -0.200 (-0.380, -0.019)* |
| e06373 | COL4A6 | 257 (94.7-433) | 0.076±0.065 | 216 (75.2-371) | -0.203±0.064 | -0.278 (-0.462, -0.945)* |

Abbreviations: ID, identification number of the sequenced peptide; IQR, interquartile range; LMean, least square mean; SE, standard error; $\Delta$ (95% CI), between-group difference (spironolactone minus control) with 95% confidence interval. Medians (IQR) were derived from the “*real world*” non-normalized non-adjusted data. LMean ± SE and $\Delta$ (95% CI) rest on the rank normalized data, which met the distribution characteristics required for running the general linear model. $\Delta$ (95% CI) provides multivariable-adjusted estimates of the between-group differences in standardized units. All models were adjusted for sex, age, BMI, eGFR, and the baseline value of the urinary marker. The other covariables included current smoking and drinking, previous ischemic heart disease and treatment at baseline and change in treatment at last follow-up with antihypertensive, lipid-lowering, antiplatelet and antidiabetic drugs. Significance of the between-group differences were adjusted for multiple comparisons by the Benjamini-Hochberg approach: \* $P\leq 0.05$ ; † $P\leq 0.01$ ; ‡ $P\leq 0.001$ (see **Table 2** for exact $P$ -values).

With control patients as reference, the differences in urinary peptides on spironolactone in HOMAGE are shown in **Table 3**. Of the 11 peptides derived from COL1A1, 7 had higher abundance on spironolactone and 4 lower abundance. Of 4 peptides derived from COL3A1, 3 were reduced and 1 increased. Of the 19 peptides derived from the collagen α1 chain, 9 were decreased; of all 27 collagen fragments, 16 were reduced. The corresponding results for PRIORITY are listed in **Table 4**. All collagen derived peptides were lowered on spironolactone with the exception of one fragment from COL1A2, one from COL5A2, and one from COL4A6. The 9 peptides derived from COL1A1 were all reduced.

**Table 4.** Between-Group Differences in the Urinary Peptide Levels at Last Follow-Up in the PRIORITY Trial Subset.

| Urinary Peptide | | Placebo (n=61) | | Spironolactone (n=49) | | $\Delta$ (95% CI) |
| --- | --- | --- | --- | --- | --- | --- |
| ID | Symbol | Median (IQR) | LMean±SE | Median (IQR) | LMean±SE | |
| e10142 | COL1A1 | 538 (347-908) | 0.299±0.132 | 310 (221-531) | -0.372±0.149 | -0.671 (-1.083, -0.260)* |
| e11829 | COL2A1 | 8.4 (8.3-8.5) | -0.168±0.104 | 8.4 (8.3-11.4) | 0.210±0.117 | -0.378 (-0.701, -0.055)* |
| e11222 | COL3A1 | 409 (165-802) | 0.259±0.123 | 255.4 (3.7-476.7) | -0.322±0.139 | -0.581 (-0.966, -0.195)* |
| e02022 | COL5A1 | 7.8 (7.7-7.9) | 0.167±0.097 | 7.8 (7.7-7.9) | -0.208±0.110 | -0.375 (-0.678, -0.073)* |
| e16281 | COL6A1 | 600 (122-1119) | 0.222±0.129 | 249 (46.4-778) | -0.276±0.146 | -0.498 (-0.901, -0.095)* |
| e12945 | COL11A1 | 273.3 (70.9-469.7) | 0.228±0.117 | 180.1 (11.4-387.1) | -0.284±0.132 | -0.512 (-0.880, -0.145)* |
| e02311 | COL15A1 | 531 (149-990) | 0.340±0.122 | 235.6 (17.7-528.8) | -0.423±0.138 | -0.763 (-1.143, -0.383)† |
| e05421 | COL16A1 | 34.4 (34.3-66.0) | 0.266±0.129 | 34.4 (30.8-45.8) | -0.331±0.146 | -0.596 (-1.001, -0.192)* |
| e18049 | COL22A1 | 2520 (479-4805) | 0.261±0.119 | 928.7 (8.1-2731.7) | -0.324±0.135 | -0.585 (-0.958, -0.212)* |
| e10651 | COL1A2 | 90.7 (11.9-252.6) | -0.192±0.101 | 126.1 (18.6-715.7) | 0.239±0.114 | 0.431 (0.116, 0.746)* |
| e09063 | COL5A2 | 17.4 (17.3-22.8) | -0.211±0.110 | 17.4 (17.3-104.3) | 0.262±0.124 | 0.473 (0.131, 0.816)* |
| e02189 | COL4A4 | 276 (141-508) | 0.252±0.123 | 174.3 (24.5-284.1) | -0.314±0.140 | -0.567 (-0.955, -0.179)* |
| e06373 | COL4A6 | 95.4 (43.9-222.4) | -0.222±0.112 | 159.4 (61.0-262.7) | 0.277±0.126 | 0.499 (0.149, 0.849)† |

Abbreviations: ID, identification number of the sequenced peptide; IQR, interquartile range; LMean, least square mean; SE, standard error; $\Delta$ (95% CI), between-group difference (spironolactone minus placebo) with 95% confidence interval. Medians (IQR) were derived from the “*real world*” non-normalized non-adjusted data. LMean ± SE and $\Delta$ (95% CI) rest on the rank normalized data, which met the distribution characteristics required for running the general linear model. $\Delta$ (95% CI) provides multivariable-adjusted estimates of the between-group differences in standardized units. All models were adjusted for sex, age, BMI, eGFR, and the baseline value of the urinary marker. The other covariables included smoking, previous cardiovascular disease, and treatment at baseline with antihypertensive and antidiabetic drugs). Significance of the between-group differences were adjusted for multiple comparisons by the Benjamini-Hochberg approach: \* $P \leq 0.05$ ; † $P \leq 0.01$ (see **Table 2** for $P$ -values).

### Serum Biomarkers

In this analytical subset of the HOMAGE data, there were no between-group differences at 1 and 9 months in CITP (**Table 5**). However, at 1 and 9 months, PICP and the PICP/CITP ratio were lower on spironolactone than control. At the 9-month time point (**Table 4**), the median serum PICP level amounted to 79.0 μg/L in control patients and 75.4 μg/L on spironolactone (spironolactone minus control difference in standardized units: −0.321; *P*=0.0007); the corresponding estimates for the PICP/CITP ratio were 21.3 and 18.3 (−0.256; *P*=0.0129). The correlations between the changes from baseline to follow-up in the urinary peptides and the corresponding changes in CITP, the serum marker reflecting degradation of mature type-1 collagen, revealed that these correlations were similar in patients randomized to control and spironolactone (**Table S4** and **Figure 1**). However, none of these correlations was significant if the COL1A1 fragments decreased from baseline to follow-up, whereas several correlations reached significance if the COL1A1 fragments increased during follow-up (**Table S4**).

**Figure 1.**
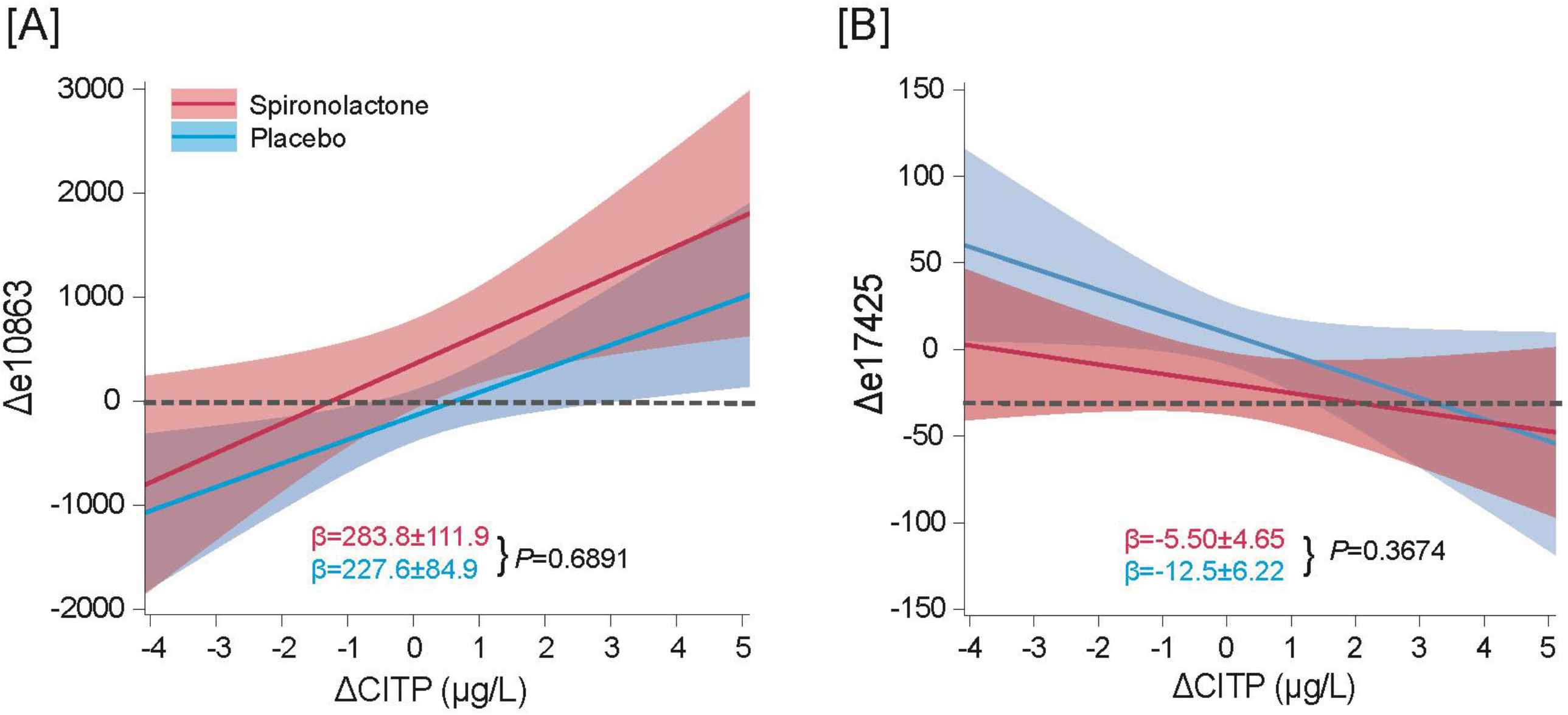
Linear Associations Between the Changes over Follow-Up (Δ) in the COL1A1-Derived Peptides e10863 (A) and e17425 (B) Regressed on the Change (Δ) in CITP. e10863 increased during follow-up, whereas e17425 decreased. CITP refers to carboxyterminal telopeptide of collagen I, a circulating marker of COL1A1 breakdown. The regression lines are presented with 95% confidence interval. The regression slopes (β) are given with standard error. The dotted lines indicate β=0. The *P*-value refers to the between-group (placebo *vs* spironolactone) in the regression slopes.

**Table 5.** Serum Fibrosis Biomarkers at Baseline and During Follow-Up in the HOMAGE Trial Subset.

| Patient Group<br>Circulating Biomarker | Placebo | | Spironolactone | | $\Delta$ (95% CI) | P-Value |
| --- | --- | --- | --- | --- | --- | --- |
| | Median (IQR) | LMean±SE | Median (IQR) | LMean±SE | | |
| PICP, μg/L | | | | | | |
| Baseline | 80.4 (66.8-96.2) | 0.022±0.084 | 80.7 (65.8-95.4) | -0.021±0.083 | -0.043 (-0.274, 0.188) | 0.7191 |
| Month 1 | 80.6 (69.3-97.2) | 0.129±0.057 | 72.9 (60.1-92.1) | -0.124±0.058 | -0.253 (-0.413, -0.093) | 0.0025 |
| Month 9 | 79.0 (66.4-100) | 0.164±0.065 | 75.4 (61.1-90.5) | -0.157±0.064 | -0.321 (-0.501, -0.142) | 0.0007 |
| CITP, μg/L | | | | | | |
| Baseline | 3.72 (2.89-5.09) | 0.029±0.082 | 3.74 (2.90-4.67) | -0.029±0.081 | -0.058 (-0.283, 0.168) | 0.6210 |
| Month 1 | 3.80 (3.10-5.08) | -0.054±0.057 | 3.93 (3.15-5.17) | 0.057±0.055 | 0.105 (-0.048, 0.257) | 0.1848 |
| Month 9 | 3.82 (3.04-4.99) | -0.038±0.065 | 4.00 (3.06-5.42) | 0.040±0.065 | 0.079 (-0.101, 0.259) | 0.4039 |
| PICP/CITP |  |  |  |  |  |  |
| Baseline | 20.8 (15.5-29.3) | 0.000±0.082 | 21.9 (16.6-28.1) | 0.001±0.081 | 0.001 (-0.225, 0.228) | 0.9912 |
| Month 1 | 22.1 (15.1-27.6) | 0.117±0.060 | 18.3 (13.8-24.6) | -0.123±0.060 | -0.240 (-0.406, -0.074) | 0.0056 |
| Month 9 | 21.3 (16.1-28.7) | 0.130±0.071 | 18.3 (13.0-26.0) | -0.126±0.070 | -0.256 (-0.451, -0.061) | 0.0129 |

Abbreviations: PICP, procollagen type I C-terminal propeptide; CITP, collagen type I C-terminal telopeptide; PICP/CITP, ratio of PICP to CITP; IQR, interquartile range; LMean, least square mean; SE, standard error; $\Delta$ (95% CI), between-group difference (spironolactone minus placebo) with 95% confidence interval; P-value, significance of the between-group differences. Medians (interquartile range) were derived from the "real world" non-normalized non-adjusted data. LMean ± SE and $\Delta$ (95% CI) rest on rank normalized data to meet the distributional characteristics required to run the general linear model. $\Delta$ (95% CI) provides multivariable-adjusted estimates of the between-group differences in standardized units. All models were adjusted for sex, age, BMI, eGFR, the baseline value of the serum biomarker, current smoking and drinking, history of ischemic heart disease, and treatment at baseline antihypertensive, lipid-lowering, antiplatelet and antidiabetic drugs. The 9-month models were additionally adjusted for the changes in the treatment status with antihypertensive, lipid-lowering, antiplatelet and antidiabetic drugs.

In the HOMAGE subset, serum sodium decreased by 0.904 mmol/L (95% CI: 0.443 to 1.364 mmol/L), whereas serum potassium increased by 0.139 mmol/L (95% CI: 0.058 to 0.219 mmol/L). Similar changes were observed in the PRIORITY subset, but only the increase serum potassium by 0.182 mmol/L (95%CI: 0.028 to 0.337 mmol/L) reached significance (**Table S5**).

## DISCUSSION

This is first report on changes in urinary peptides in response to MRA by spironolactone based on analytical subsets of the HOMAGE^3^ and PRIORITY^12^ trials. In HOMAGE (**Table 3**) and PRIORITY (**Table 4**) combined, 26/39 (66.7%) fragments from mature collagens, 6 originating from type-1 collagen, had lower abundance on spironolactone than placebo. Given the significant decrease in PICP and the PICP/CITP ratio in the HOMAGE trial subset and the stochiometric ratio relating PICP to collagen synthesis (**Figure S1**), the lower abundance on spironolactone of peptides derived from type-1 collagen might indicate a reduced synthesis. However, in the HOMAGE subset, 7 urinary fragments of COLA1A were increased on spironolactone (**Table 3**). Differences between the alterations in the urinary peptides in HOMAGE and PRIORITY probably reflect divergent pathogenic processes affecting the main target organs in patients at risk of HF and normoalbuminuric T2DM patients with normal renal function as well as the 10-year difference in calendar age (median age: 73 *vs* 64 years) with possible repercussions on biological aging.^6^ In the HOMAGE trial subset, the correlations between the changes from baseline to follow-up in the urinary peptides and in CITP, the serum marker reflecting degradation of mature type-1 collagen, revealed that the slopes of these associations were similar in patients randomized to control and spironolactone (**Figure 1** and **Table S4**). However, none of these correlations was significant if the type-1 collagen fragments decreased from baseline to follow-up, whereas several correlations reached significance if the COL1A1 fragments increased during follow-up (**Table S4**). Overall, the most likely explanation of the combined HOMAGE and PRIORITY findings is a decrease on spironolactone of the body-wide pool of collagens available for degradation leading to the lower abundance of collagen-derived urinary peptides. However, the increase on spironolactone of 7 COL1A1 fragments in relation to CITP (**Table 3** and **Table S4**) in HOMAGE also indicates that the breakdown of type-1 collagen remains unaffected.

The proposed interpretation of the current findings is in keeping with the literature. In a random-effect meta-analysis^23^ of 1038 patients randomized in HOMAGE (47.0%),^3^ ALDO-DHF (37.2%)^24^ and TOPCAT (15.7%),^25^ administration of spironolactone for 9 to 12 months compared to placebo or usual care reduced PICP by 7.4 μg/L (95% CI: 0.9 to 13.9 μg/L). This association between spironolactone and serum PICP was not mediated by BP.^23^ This meta-analysis was consistent with the concept that spironolactone reduces type-1 collagens in patients with stages 3-4 HF.^23^ In a post-hoc analysis of 1411 patients receiving spironolactone as add-on therapy in the ASCOT-BPLA, the serum concentrations of PICP and PIIINP rose in controls and fell on spironolactone treatment. The adjusted mean changes were +0.52 (95% CI: −0.05 to 1.09) *vs* −0.41 (−0.97 to 0.16) μg/L for PIIINP and +4.54 (−1.77 to 10.9) *vs* −6.36 (−12.5 to −0.21) μg/L; *P*=0.023 for PICP.^26^ An aptamer-based proteomic analysis utilized 5284 modified aptamers to 4928 unique proteins in 164 TOPCAT patients with paired plasma samples at baseline and 1 year.^27^ The top 4 canonical pathways were enriched for multiple collagens that increased in the placebo group, but decreased with spironolactone.^27^ CARD18 was the most upregulated protein on spironolactone.^27^ CARD18 is a small caspase recruitment domain-containing decoy molecule induced by proinflammatory stimuli but inhibits caspase-1 oligomerization and activation and subsequent generation of IL (interleukin)-1β.^28^

A recent study examined 5000 UPP datasets randomly selected from healthy participants and patients with CKD.^9^ Well-correlated peptides with the same amino-acid sequence, but a different number of hydroxyprolines, were combined into a final list of 503 peptides, which covered 69% of the full COL1A1 sequence. Sixty-three COL1A1 fragments (12.5%) were positively correlated (*r* > 0.30) with eGFR, suggesting that dysfunction of the kidney, and potentially of other organs, might be due to an attenuation of collagen degradation.^9^ However, studies which reported a decrease in urinary peptide fragments derived from COL1A1 as the hallmark of fibrosis,^7-9^ had an observational design. Such studies do not allow to draw conclusions about active intervention with MRAs, such as in the context of randomized clinical trials of spironolactone.

Injury activates resident fibroblasts or mobilizes bone-marrow-derived circulating fibrocytes and epithelial or endothelial cells, and drives the epithelial/endothelial-to-mesenchymal transition.^29^ Subsequently, these cells transdifferentiate into α-smooth muscle actin-expressing myofibroblasts that secrete the ECM components required for wound repair in acute injury, but produce excessive ECM deposition in response to persistent injury.^29^ Antifibrotic drugs remain a critically important unmet medical need, as nearly 45% of all natural deaths in the Western world are attributable to the complications of chronic fibroproliferative disorders.^30^ A detailed review of the molecular pathways of fibrosis and of the refurbished or experimental drugs inhibiting fibrosis extra- or intra-cellularly is beyond the scope of this article but was summarized elsewhere.^31^ However, drugs specifically designed to inhibit fibrosis at the intra- or extra-cellular level, in particular by activating collagen degradation, would meet an imperative need in aging populations,^30^ but few are being actively developed.^31^ The underlying reason might be that the current ontology of disease focuses on specific organs rather than on the generalized pathophysiological processes, fibrosis being a major bodywide actor, that transcends disease classifications. Because our findings suggest that collagen deposition exceeds its breakdown, focus might shift to pharmacological interventions promoting resolution of fibrosis in addition to inhibition of its deposition. This concept would strengthen the therapeutic armamentarium to modify fibrosis, but is also challenging, because mature collagen is a key factor in the healing process in response to acute injury. Interventions targeting metalloproteinases had off-target adverse effects.^31,32^ On the other hand, nonsteroidal MR antagonists and sodium-glucose cotransporter-2 inhibitors have potent anti-inflammatory and antifibrotic properties. UPP might open new perspectives in documenting the antifibrotic properties of these novel drug classes.

The present study has limitations. First, in the analytical subset of the HOMAGE data, changes in CITP were not significant, because of the smaller sample size compared with the full trial,^3^ albeit that trends were similar. Second, one possible drawback of the CE-MS approach is the application of the ultrafiltration with the threshold set at 20 kDa, so that larger proteins escape analysis. Second, proteases active along the nephron and distal urinary tract might affect the urinary peptide fragments detected by UPP analysis. However, in a placebo-controlled study of a dipeptidyl peptidase-4 inhibitor,^33^ the UPP included pairs of peptide chains, i.e., the substrate for the protease activity (e.g., PPGPPGKNGDDGEAGKPG) and the resulting breakdown product (e.g., GPPGKNGDDGEAGKPG). In the current study, the UPP did not contain such peptide pairs, so that the assumption that MRA influenced the UPP by changing protease activity along the urinary tract can be disregarded. Finally, CITP is the small fragment released from type-1 collagen during the first step in its degradation mediated by collagenase (**Figure S1**). The larger fragment is further degraded by gelatinases with resulting smaller fragments that are further degraded by other metalloproteinases. Thus, the peptides measured in urine are likely those released by gelatinases and those subsequently released by metalloproteinases. These fragments, named matricryptins, may have biological activity, which contribute to the regulation of inflammatory, reparative, and fibrogenic cascades, but for which the current analyses did not account for.

## CONCLUSIONS

MRA by spironolactone reduces collagen-derived urinary peptides. Inhibition of collagen synthesis reducing the amount available for breakdown may be a contributing mechanism. MRA does not affect the relation between urinary peptides and serum CITP, a marker of collagen degradation.

### Conflict of Interests

Justyna Siwy and Agnieszka Latosinska are employees of Mosaiques-Diagnostics GmbH, Hannover, Germany. Harald Mischak is the co-founder and co-owner of Mosaiques-Diagnostics GmbH. The other authors declare no conflict of interest.

## Funding

PRIORITY and HOMAGE were funded by the European Union Seventh Framework Program. OMRON Healthcare, Co., Ltd., Kyoto, Japan provided a non-binding grant to the Non-Profit Research Association Alliance for the Promotion of Preventive Medicine (APPREMED), Mechelen, Belgium.

## Data Availability

All data produced in the present study are available upon reasonable request to the authors.

## Abbreviations and Acronyms

95% CI: 95% confidence interval
Aldo-DHF: the Aldo-DHF Randomized Controlled Trial
BNP: brain natriuretic peptide
BMI: body mass index
BP: blood pressure
CE-MS: capillary electrophoresis coupled with mass spectrometry
CITP: carboxyterminal telopeptide of collagen I
CKD: chronic kidney disease
Δ: change from baseline to last follow-up
eGFR: glomerular filtration rate derived from serum creatinine by the Chronic Kidney Disease Epidemiology (CKD-EPI) formula
ECM: extracellular matrix
HF: heart failure
HOMAGE: Omics in Ageing Randomized Clinical Trial
MOS: Mosaiques-Diagnostiques GmbH, Hannover, Germany
MR: mineralocorticosteroid receptor
MRA: mineralocorticoid receptor antagonist
NTproBNP: amino-terminal pro-B-type natriuretic peptide
PICP: carboxyterminal propeptide of procollagen I
PIIINP: amino terminal propeptide of procollagen type III
PRIORITY: Proteomic Prediction and Renin Angiotensin Aldosterone System Inhibition Prevention of Early Diabetic Nephropathy in Type 2 Diabetic Patients with Normoalbuminuria
TOPCAT: Treatment of Preserved Cardiac Function Heart Failure with an Aldosterone Antagonist
UPP: urinary proteomic profile

## Acknowledgements

The authors are indebted to the many investigators, who were involved in HOMAGE and PRIORITY. Their names are listed in the Data Supplement. This article was submitted for publication on their behalf.

## Notes

### Clinical Trial

NCT02556450; NCT02040441

### Clinical Protocols

https://academic.oup.com/eurheartj/article/42/6/684/5993916?login=false

https://pubmed.ncbi.nlm.nih.gov/32135136/

### Author Declarations

Ethics committees of the following institutions gave ethical approval for this work. The HOMAGE trial was in 9 centers in the United Kingdom, France, Italy, Ireland, Germany and the Netherlands. Patients were screened in primary and secondary care. Each center had its own recruitment strategies under review of local ethics committees (see statistical analysis plan available at www.clinicaltrials.gov). The PRIORITY study was conducted at 15 specialist centers in 10 European countries (Belgium, Czech Republic, Denmark, Germany, Greece, Italy, the Netherlands, North Macedonia, Spain, and the UK; a lists of sites and the local ethics approval numbers are available in reference 12. (Ethics committees include Region Hovedstaden Regionsgården Kongens, Sächsische Landesärztekammer, Commissie voor Medische Ethiek Universitair Ziekenhuis, Ethics Committee of the Institute for Clinical and Experimental Medicine and Thomayer Hospital, Ethics Committee of the University Hospital Kralovske Vinohrady, CEIC Fundación Jiménez Díaz, EAAHNIKH AHMOKPATIA YIIOYPTEIO YTEIAΣ (NEC) EAAHNIKH AHMOKPATIA 1n Y.H.E. ATTIKHE rEN.NOI/MEIO A0HNAI inflOKPATElO (HEC), Comitato Etico della Provincia di Bergamo, Medisch Ethische Toetsingscommissie, Universitair Medish Centrum Groningen, West of Scotland REC 1 R&D Management Office Western Infirmary Tennent Institute R&D)

