## Supplementary material for "Urinary Proteomic Signature of Mineralocorticoid Receptor Antagonism by Spironolactone: Evidence from the Randomized-Controlled HOMAGE and PRIORITY Trials": Table S1; Table S2; Table S3; Table S4; Table S5; Figure S1; Figure S2; Figure S3; Figure S4; Figure S5

#### Table of Contents

**HOMAGE Investigators** p2

**PRIORITY Investigators** p3

**Urinary Proteomics** p4

Sample preparation and CE-MS analysis p4

CE-MS data processing p4

Sequencing of peptides p5

References p6

**Serum Biomarkers** p7

**FIGURE S1** Circulating Biomarkers of Collagen Turnover p8

**FIGURE S2** Flow Chart of the HOMAGE Analytical Subset p9

**FIGURE S3** Flow Chart of the PRIORITY Analytical Subset p10

**FIGURE S4** Rank Normalization of CITP as Marker of Collagen Degradation p11

**FIGURE S5** Rank Normalization of Urinary Peptide Fragment e08916 Derived from Collagen 1 p12

**TABLE S1** Comparison of Patients Analyzed and Not Analyzed p13

**TABLE S2** Urinary Peptide Sequences Retained in the Analyses p15

**TABLE S3** Comparison of the Baseline Urinary Peptide Levels Between Randomized Groups p18

**TABLE S4** Correlations Between the Changes over Follow-up in COL1A1-Derived Urinary Peptides and in CITP in the HOMAGE Subset p20

**TABLE S5** Between-Group Differences in Serum Electrolytes and Renal Function at Last Follow-Up p21

### HOMAGE Investigators

*Robertson Centre for Biostatistics, Institute of Health and Wellbeing, University of Glasgow, Glasgow, Scotland, UK* — John GF Cleland, MD; Pierpaolo Pellicori, MD; Javed Khan, MD.

*Université de Lorraine, Inserm, Centre d'Investigation Clinique Plurithématique, CHRU de Nancy, Nancy, France* — João P Ferreira, MD; Franco Cosmi, MD; Anne Pizard, PhD; Nicolas Girerd, MD; Patrick Rossignol, MD; Erwan Bozec, MD; María U Moreno, MD; Faiez Zannad, MD.

*Department of Cardiology, Cortona Hospital, Arezzo, Italy* — Beatrice Mariottoni, MD.

*Department of Cardiology, University of Hull, Castle Hill Hospital, Cottingham, East Riding of Yorkshire, UK* — Joe Cuthbert, MD; Andrew L Clark, MD.

*Department of Cardiology, Maastricht University Medical Center, Maastricht, the Netherlands* — Job AJ Verdonschot PhD; Hans P Brunner La Rocca, MD; Mark Hazebroek, PhD; Stephane Heymans, MD.

*Department of Internal Medicine and Cardiology, Campus Virchow Klinikum, Charité, University Medicine Berlin, Berlin Institute of Health (BIH), and German Centre for Cardiovascular research (DZHK), Partner Site Berlin, Germany* — Johannes Petutschnigg, MD; Frank Edelmann, MD; Burkert Pieske, MD.

*Division of Cardiovascular Sciences, School of Medical Sciences, Faculty of Biology, Medicine and Health, Manchester Academic Health Science Centre, University of Manchester, Manchester, UK* — Fozia Z Ahmed, MD; Mamas A Mamas, MD.

*Centre for Prognosis Research, Institute for Primary Care and Health Sciences, Keele University, Newcastle, UK* — Mamas A Mamas, MD.

*German Heart Center Berlin, Berlin, Germany* — Burkert Pieske, MD.

*St. Vincent's University Healthcare Group, and School of Medicine, University College Dublin, Dublin, Ireland* — Ken McDonald, MD.

*Equipe obésité et insuffisance cardiaque, Université Paul Sabatier, Inserm I2MC, Toulouse, France* — Philippe Rouet, MD.

*Studies Coordinating Centre, Research Unit Hypertension and Cardiovascular Epidemiology, Department of Cardiovascular Sciences, University of Leuven, Leuven, Belgium* — L Thijs, MSc.

*Non-Profit Research Association Alliance for the Promotion of Preventive Medicine, Mechelen, Belgium* — Jan A Staessen, MD; Kei Asayama, MD; Tine W Hansen, MD; Gladys E Maestre, MD.

*Program of Cardiovascular Diseases, CIMA. Universidad de Navarra and IdiSNA, Pamplona, Spain CIBERCV, Carlos III Institute of Health, Madrid, Spain* — Arantxa González, PhD; Suzanna Ravassa, PhD; Begoñia López, PhD; Javier Díez, MD.

*Departments of Nephrology and Cardiology, Clínica Universidad de Navarra, Pamplona, Spain* — Javier Díez, MD.

*Department of Cardiovascular Medicine, Istituto di Ricerche Farmacologiche Mario Negri – IRCCS, Milan, Italy* — Roberto Latini, MD.

*Fondation Force, Research and Consulting Department, EDDH, Centre de Médecine Préventive, Vandoeuvre les Nancy, France* — Stephanie Grojean, PhD.

*Department of Medical Statistics, London School of Hygiene and Tropical Medicine, London, UK* — Tim Collier, PhD.

### PRIORITY Investigators

*Steno Diabetes Center Copenhagen, Copenhagen, Denmark* — Peter Rossing, MD; Morten K Lindhardt, MD; Nete Tofte, MD; Marie Frimodt-Møller, MD; Viktor Rotbain Curovic, MD; Frederik Person, MD.

*Mosaiques-Diagnostics GmbH, Hannover, Germany* — Harald Mischak, PhD; Petra Zürbig, PhD; Justyna Siwy, PhD.

*Hannover Clinical Trial Center, Hannover Medical School, Hannover, Germany* — Heiko van der Leyen, PhD.

*University Medical Center Groningen, Groningen, the Netherlands*—Gerjan Navis, MD; Stephan JL Bakker, MD; Gozewijn D Laverman, MD; Hiddo JL Heerspink, PhD.

*NHS Greater Glasgow and Clyde Clinical Research Facility, Glasgow UK* — Joanne Flynn, MD.

*University of Glasgow, Glasgow, UK* — Christian Delles, MD; Gemma Currie, MD; John R Petrie, MD.

*Istituto di Ricerche Farmacologiche Mario Negri, Bergamo Italy* — Pierro L Ruggenenti, MD; Matias Trillini, MD; A Parvanova, MD.

*Charles University Prague, Prague, Czech Republic* — Ivan Rychlik, MD; Lidmila Francová, MD.

*National and Kapodistrian University of Athens, Athens, Greece* — Marina Noutsou, MD.

*Institute for Clinical and Experimental Medicine Prague, Prague, Czech Republic* — Peter Girman, MD; Tereza Havrdova, MD.

*Instituto de Investigacion, Sanitaria de la Fundacion Jiménez Díaz, Madrid, Spain* — Alberto Ortiz, MD; Beatrice Fernandez-Fernandez, MD.

*Hospital St. Georg, Leipzig, Germany* — Joachim Beige, MD; Ingo Dimos, MD.

*University Clinic of Endocrinology, Skopje, North Macedonia* — Goce Spasovski, MD; Katarina Adamova, MD.

*Bethesda Diabetes Research Center, Hoogeveen, the Netherlands* — Adriaan Kooy, MD.

*Ghent University Hospital, Ghent, Belgium* — Marijn Speeckaert, MD.

*University Hospital Tübingen, Tübingen, Germany* — Andreas Birkenfeld, MD.

*VUMC Amsterdam, Amsterdam, the Netherlands* — Jolien WJ Beulens, MD; Femke Rutters, MD; Giel Nijpels, MD.

*Diabetologen Hessen, Hessen, Germany*—Rüdiger Göke, MD.

### Urinary Proteomics

All steps of the CE-MS analysis and the performance of the analytical platform have recently been described in detail.[1]

#### *Sample Preparation and CE-MS Analysis*

Urine aliquots were thawed and 700 µL mixed with 700 µl of 2 M urea, 10 mM $NH_4OH$ containing 0·02 % SDS. Subsequently, samples were ultrafiltered using a Centristat 20 kDa cut-off centrifugal filter device (Satorius, Göttingen, Germany) to eliminate high molecular weight proteins. The obtained filtrate was desalted using a PD 10 gel filtration column (GE Healthcare Bio Sciences, Uppsala, Sweden) to remove urea, electrolytes and salts as well as to enrich polypeptides. The samples were lyophilized and stored at 4°C before usage. Shortly before CE-MS analysis, the samples were re-suspended in 10 µL HPLC-grade $H_2O$. Samples were injected into CE-MS with 2 psi for 99 sec, resulting in injection volumes of ~280 nL.

A P/ACE MDQ capillary electrophoresis system (Beckman Coulter, Fullerton, CA) was coupled with a Micro-TOF MS (Bruker Daltronic, Bremen, Germany). A solution of 20% acetonitrile (Sigma-Aldrich, Taufkirchen, Germany) in HPLC-grade water (Roth, Karlsruhe, Germany) supplemented with 0·94% formic acid (Sigma-Aldrich) was used as running buffer. For CE-MS analysis, the electrospray ionization interface from Agilent Technologies (Palo Alto, CA) was set to a potential of -4·0 to -4·5 kV. Spectra were recorded over an *m*/*z* range of 350-3000 and accumulated every 3 s.

#### *CE-MS Data Processing*

After the CE-MS analysis, mass spectral ion peaks representing identical molecules at different charge states were deconvoluted into single masses using MosaFinder software.[2] Only signals with z>1 observed in a minimum of 3 consecutive spectra with a signal-to-noise ratio of at least 4 were considered. The resulting peak list characterizes each polypeptide by its mass and migration time. Data were calibrated utilizing 3151 internal standards as reference data points for mass and migration time by applying global and local linear regression, respectively. Reference signals of 29 abundant peptides were used as internal standards for calibration of signal intensity using linear regression. This procedure is highly reproducible and addresses both analytical and dilution variances in a single calibration step.[3] The obtained peak list characterizes each polypeptide by its calibrated molecular mass [Da], calibrated CE migration time [min] and normalized signal intensity. All detected peptides are deposited, matched, and annotated in a Microsoft SQL database allowing further statistical analysis.

***Sequencing of Peptides***

Candidate biomarkers were sequenced using CE-MS/MS or LC-MS/MS analysis, as described in detail.[4] MS/MS experiments were using an Ultimate 3000 nano-flow system (Dionex/LC Packings, USA) or a P/ACE MDQ capillary electrophoresis system (Beckman Coulter, Fullerton, CA), both connected to an LTQ Orbitrap hybrid mass spectrometer (Thermo Fisher Scientific, Germany) equipped with a nano-electrospray ion source. The mass spectrometer is operated in data-dependent mode to automatically switch between MS and MS/MS acquisition. Survey full-scan MS spectra (from $m/z$ 300–2,000) were acquired in the Orbitrap. Ions were sequentially isolated for fragmentation. Data files were searched against the UniProt human nonredundant database using Proteome Discoverer 2·4 and the SEQUEST search engine. Relevant settings were: no fixed modifications, oxidation of methionine and proline as variable modifications. The minimum precursor mass was set to 790 Da, maximum precursor mass to 6000 Da with a minimum peak count of 10. The high-confidence peptides were defined by cross-correlation (Xcorr) >1.9 and rank = 1. Precursor mass tolerance and fragment mass tolerance were 5 ppm and 0.05 Da, respectively. For further validation of obtained peptide sequences, the correlation between peptide charge at the working pH of 2 and CE-migration time was utilized to minimize incorrect sequence assignment:[5] calculated CE-migration time of the sequence candidate based on its peptide sequence (number of basic amino acids) was compared to the experimental migration time.

#### Serum Biomarkers

PICP was quantified by an enzyme linked immunosorbent assay (Quidel Corporation, San Diego, CA) and CITP by a quantitative radio-immunoassay (Orion Diagnostica, Espoo, Finland). The detection limits were 0.2 μg/L for PICP and 0.6 μg/L for CITP. All inter- and intra-assay coefficients of variation were <10%.

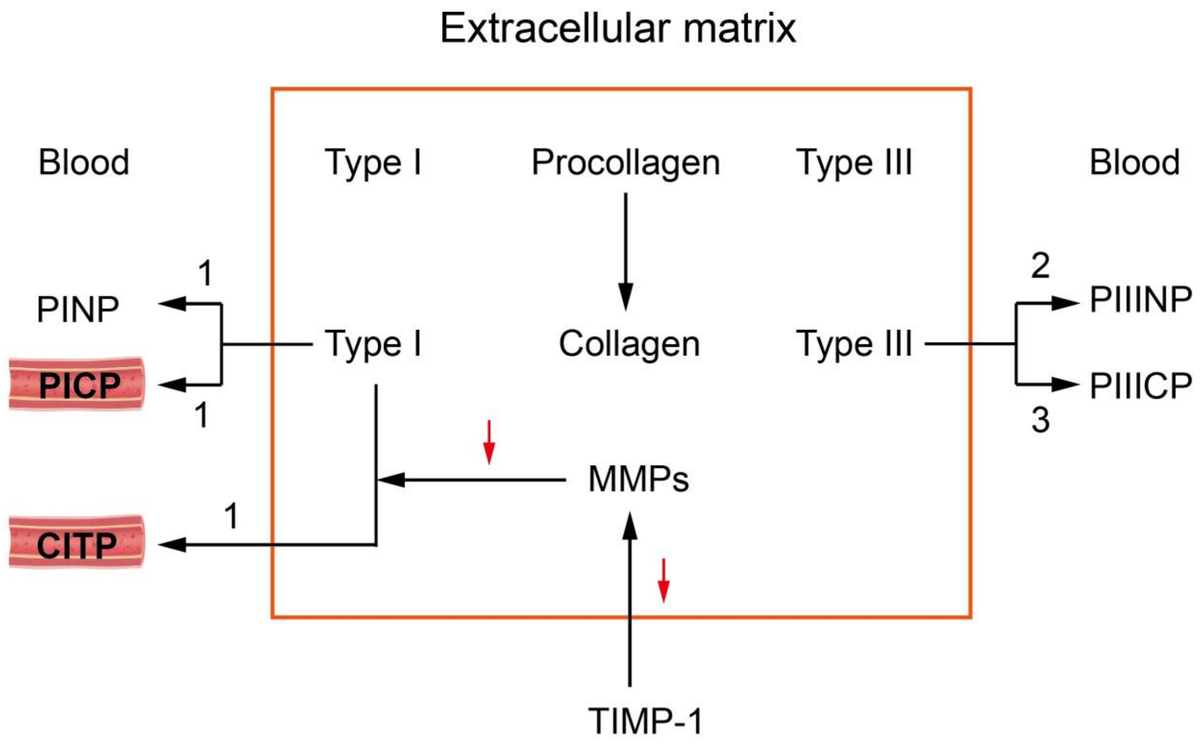


**Figure S1**

**Circulating Biomarkers of Collagen Turnover.**

PINP and PICP are released during conversion of procollagen type-1 to collagen type-1 and CITP during the degradation of collagen type-1 by matrix metalloproteinases, which are inhibited by TIMP1. PIIINP and PIIICP are released during conversion of procollagen type-III to collagen type-III. Bracketed numbers indicate the stoichiometric ratio. PICP and CITP, which were analyzed in the current HOMAGE Trial subset, are serum markers of collagen type-1 synthesis and degradation, respectively. PIIINP is an indirect indicator of collagen-III synthesis, because cleavage at the amino-terminus proceeds at a relatively slow rate and, thus, partially processed procollagen molecules remain bound to the surface of collagen type-III fibers (*JACC* 2015;65:2449-2456). Abbreviations: PICP, procollagen type-I carboxy-terminal propeptide; PINP, procollagen type-I amino-terminal propeptide; CITP, carboxyterminal telopeptide of type-I collagen; MMPs, matrix metalloproteinases; TIMP1, tissue inhibitor of the matrix metalloproteinase type-1; PIIICP, procollagen type-III carboxy-terminal propeptide; and PIIINP, procollagen type-III amino-terminal propeptide.

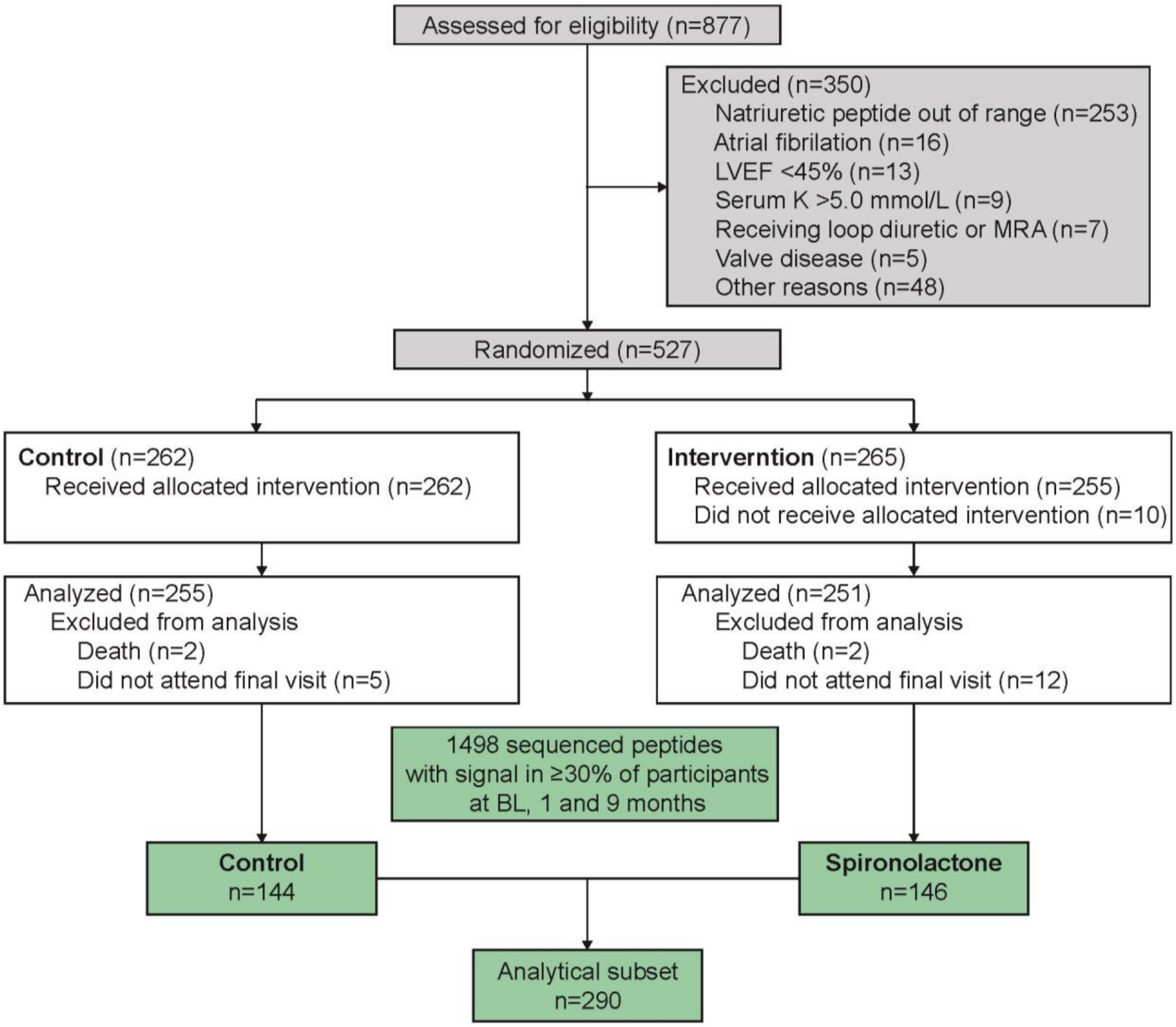


**Figure S2**

**Flow Chart of the HOMAGE Analytical Subset.**

Abbreviations: LVEF, left ventricular ejection fraction; BL, baseline.

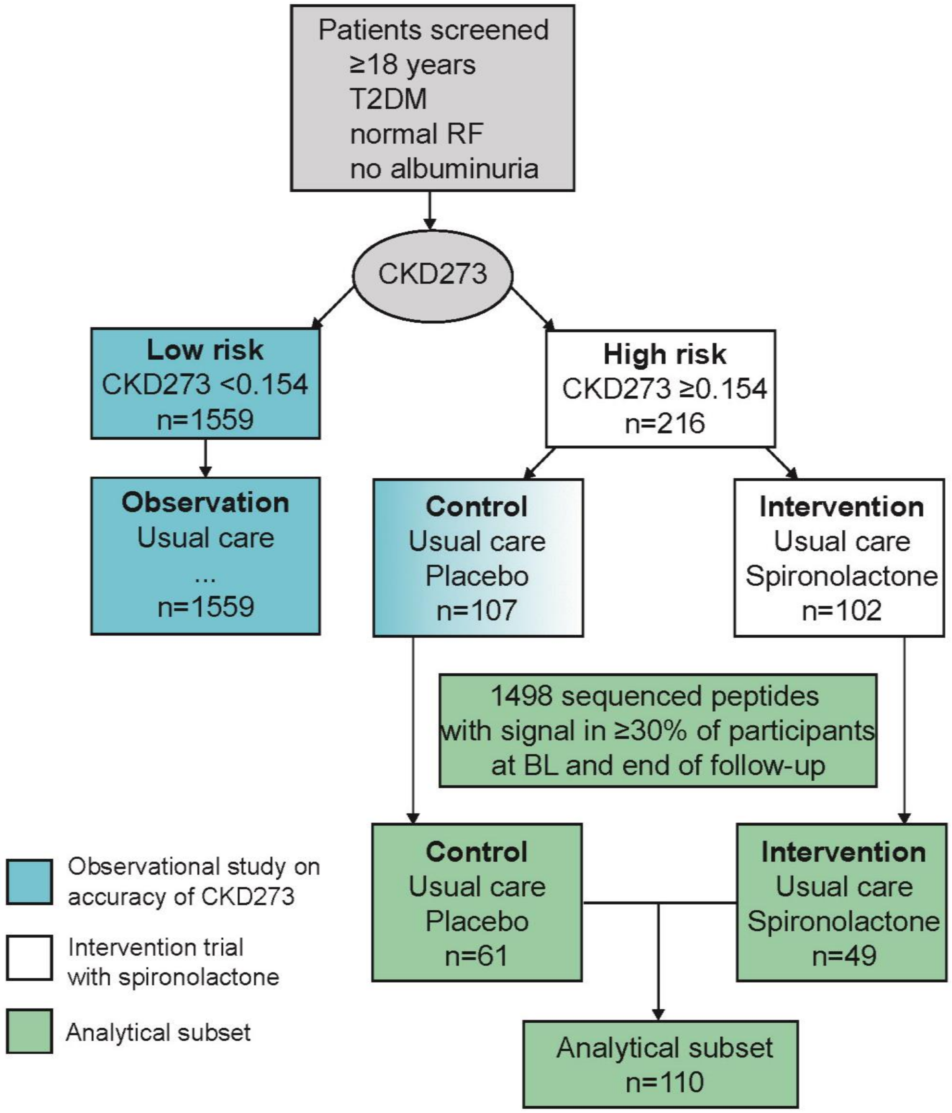


**Figure S3**

**Flow Chart of the PRIORITY Analytical Subset.**

Abbreviation: T2DM, type-2 diabetes; BL, baseline.

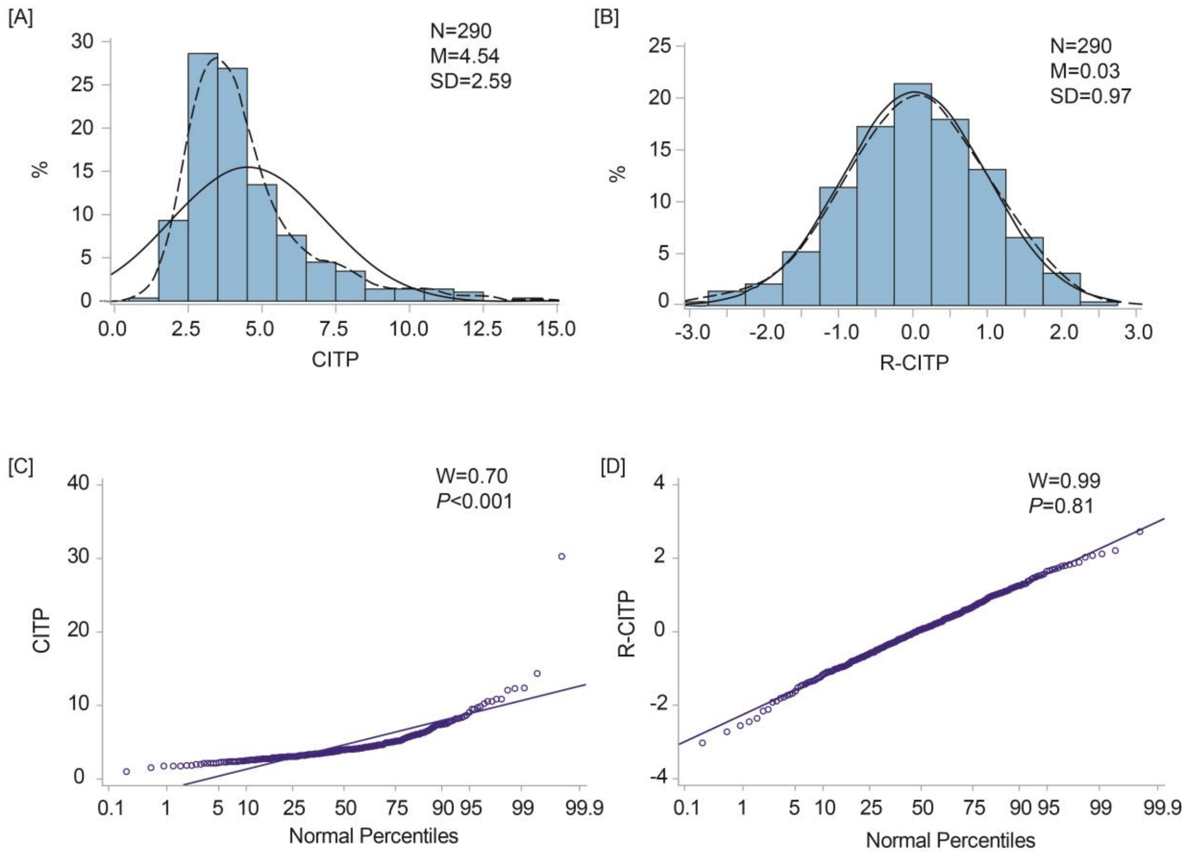


**Figure S4**

**Rank Normalization of a Carboxyterminal Telopeptide of Collagen I (CITP), a Serum Marker of Collagen I Degradation.**

Panels A and B show the distribution plots before (A) and after (B) rank-normalization; panels C and D show the normal percentile plots before (C) and after (D) rank normalization. The solid and dotted lines represent the normal and kernel density distributions. N, M and SD refer to the number of patients, the arithmetic means and standard deviation. W is the Shapiro-Wilk statistic and *P* is the associated significance. A significant Shapiro-Wilk test indicates deviation from the normal distribution.

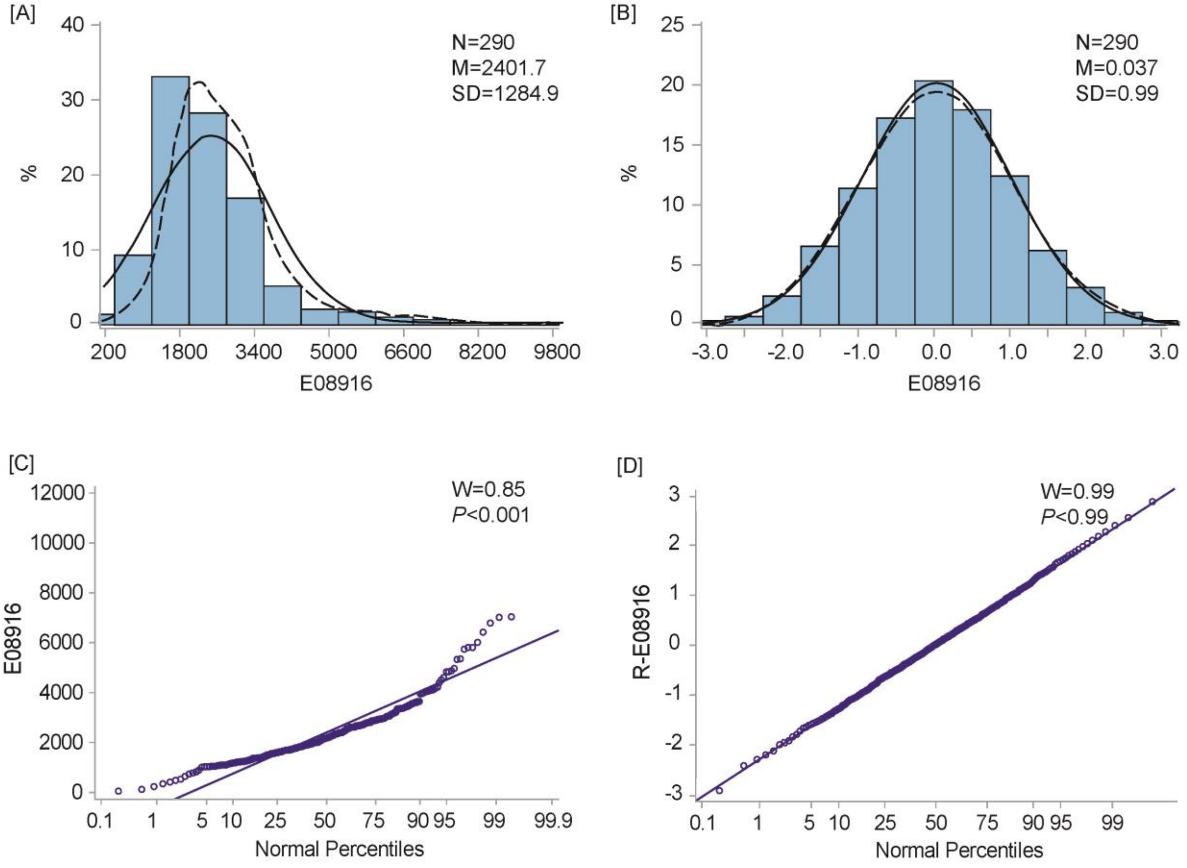


**Figure S5**

**Rank Normalization of Urinary Peptide Fragment E08916 Derived from Collagen 1.**

Panels A and B show the distribution plots before (A) and after (B) rank-normalization; panels C and D show the normal percentile plots before (C) and after (D) rank normalization. The solid and dotted lines represent the normal and kernel density distributions. N, M and SD refer to the number of patients, the arithmetic means and standard deviation. W is the Shapiro-Wilk statistic and *P* is the associated significance. A significant Shapiro-Wilk test indicates deviation from the normal distribution.

**Table S1 Comparison of Patients Analyzed and Not Analyzed (Starts)**

| **Characteristic** | **HOMAGE-RCT** | | | **PRIORITY-RCT** | | |
|---|---|---|---|---|---|---|
| | **Analyzed** | **Not analyzed** | ***P*** | **Analyzed** | **Not Analyzed** | ***P*** |
| **Number in group** | 290 | 215 | … | 110 | 99 | … |
| **Number (%)** | | | | | | |
| Women | 69 (23.8) | 57 (26.5) | 0.4851 | 24 (21.8) | 38 (38.4) | 0.0088 |
| Past smoking | 171 (59.0) | 117 (54.4) | | 45 (40.9) | 32 (32.3) | 0.0504 |
| Current smoking | 19 (6.5) | 23 (10.7) | 0.0693 | 7 (6.4) | 12 (12.1) | 0.0687 |
| Drinking alcohol | 69 (23.8) | 20 (9.3) | <0.0001 | 0 (0.0) | 0 (0.0) | ... |
| Hypertension | 217 (74.8) | 177 (82.3) | 0.0442 | 98 (89.1) | 88 (88.9) | 0.9628 |
| Diabetes | 64 (22.1) | 20 (9.3) | 0.0001 | 100 (100.0) | 99 (100.0) | ... |
| Antihypertensive treatment | 280 (96.6) | 92 (42.8) | 0.1096 | 102 (92.7) | 89 (89.9) | 0.9807 |
| Lipid-lowering treatment | 257 (88.6) | 159 (74.0) | <0.0001 | 93 (84.6) | 57 (57.6) | <0.0001 |
| Antiplatelet drugs | 218 (75.5) | 142 (66.0) | 0.0250 | 66 (60.0) | 37 (37.4) | 0.0040 |
| Antidiabetic agents | 257 (88.6) | 201 (93.5) | 0.1337 | 107 (97.3) | 87 (87.9) | 0.0086 |
| History of CVD | ... (81.0) | ... | ... | 30 (27.3) | 20 (20.2) | 0.2316 |
| History of IHD | 235 (81.1) | 130 (60.5) | <0.0001 | 23 (20.9) | 14 (14.1) | 0.2006 |
| **Clinical characteristics** | | | | | | |
| Age, y | 73.8±6.0 | 73.5±7.3 | 0.5879 | 63.2±6.5 | 63.5±6.3 | 0.7506 |
| BMI, kg/m$^2$ | 28.8±5.1 | 28.9±5.0 | 0.7618 | 30.5±5.5 | 31.3±5.4 | 0.3864 |
| Systolic BP, mm Hg | 138.3±18.2 | 139.5±17.7 | 0.4758 | 133.7±13.0 | 135.7±10.8 | 0.2244 |

**Table S1 Comparison of Patients Analyzed and Not Analyzed (Ends)**

| Characteristic | HOMAGE-RCT | | | PRIORITY-RCT | | |
|---|---|---|---|---|---|---|
| | Analyzed | Not Analyzed | *P* | Analyzed | Not Analyzed | *P* |
| Diastolic BP, mm Hg | 77.6±10.3 | 77.8±9.6 | 0.8491 | 77.2±8.7 | 80.3±9.1 | 0.0124 |
| Heart rate, bpm | 61.4±8.7 | 62.1±8.5 | 0.4201 | 76.3±13.5 | 74.6±11.4 | 0.2187 |
| **Routine biochemistry** | | | | | | |
| Serum sodium, mmol/L | 139.2±2.6 | 139.1±2.6 | 0.5734 | 139.5±2.6 | 139.7±2.6 | 0.7050 |
| Serum potassium, mmol/L | 4.36±0.37 | 4.33±0.33 | 0.3787 | 4.23±0.44 | 4.26±0.42 | 0.5972 |
| eGFR, mL/min/1.73 $m^2$ | 67.1±15.3 | 70.8±17.4 | 0.0122 | 81.9±16.8 | 80.7±17.1 | 0.6151 |
| Total cholesterol, mg/dL | 149.8±42.4 | 141.4±53.1 | 0.0479 | 162.2±43.9 | 176.7±40.3 | 0.0226 |
| HbA1C, % | 6.11±1.31 | 6.13±1.02 | 0.8979 | 8.02±1.91 | 7.83±1.60 | 0.4389 |
| UACR, mg/g | ... | ... | ... | 8.29 (5.03-12.17) | 5.58 (3.79-10.24) | 0.0449 |

Abbreviations: CVD, cardiovascular disease; IHD, ischemic heart disease; BP, blood pressure; BMI, body mass index; eGFR, glomerular filtration rate estimated from serum creatinine according to the Chronic Kidney Disease Epidemiology equation; HBA1C, glycated hemoglobin; UACR, urinary albumin-to-creatinine ratio. Tabulated values are number of patients (%), arithmetic mean ± SD, or median (IQR) for variables deviating from the normal distribution. *P*-values were computed by the large sample Z test (after rank normalization, if appropriate) for continuously distributed variables or by the $\chi^2$ statistic or Fisher's exact test, as appropriate according to the cell frequencies, for categorical variables. An ellipsis indicates that the variable was not measured.

**Table S2 Urinary Peptide Sequences Retained in the Analyses (Starts)**

| **ID** | **Symbol** | **Amino-Acid Sequence** | **Protein (*Uniprot ID*)** |
|---|---|---|---|
| **e04960** | COL1A1 | GppGpPGSpGEQGPSG | collagen alpha-1 (I) chain (*P02452)* |
| **e08916** | COL1A1 | DDGEAGKPGRpGERGpPGP | collagen alpha-1 (I) chain (*P02452*) |
| **e09408** | COL1A1 | GDDGEAGKpGRPGERGPPGp | collagen alpha-1 (I) chain (*P02452*) |
| **e10142** | COL1A1 | EGSpGRDGSpGAKGDRGETGp | collagen alpha-1 (I) chain (*P02452*) |
| **e10266** | COL1A1 | NGDDGEAGKpGRpGERGPPGP | collagen alpha-1 (I) chain (*P02452*) |
| **e10395** | COL1A1 | NGDDGEAGKpGRPGERGpPGp | collagen alpha-1 (I) chain (*P02452*) |
| **e10437** | COL1A1 | AEGSPGRDGSpGAKGDRGETGP | collagen alpha-1 (I) chain (*P02452*) |
| **e10657** | COL1A1 | AEGSpGRDGSpGAKGDRGETGp | collagen alpha-1 (I) chain (*P02452*) |
| **e10863** | COL1A1 | DGQPGAKGEpGDAGAKGDAGPPGp | collagen alpha-1 (I) chain (*P02452*) |
| **e11972** | COL1A1 | ADGQpGAKGEPGDAGAKGDAGppGPA | collagen alpha-1 (I) chain (*P02452*) |
| **e17425** | COL1A1 | AGPTGARGAPGDRGEPGPpGpAGFAGpPGADGQPGAK | collagen alpha-1 (I) chain (*P02452*) |
| **e18740** | COL1A1 | DKGETGEQGDRGIKGHRGFSGLQGppGPPGSPGEQGP | collagen alpha-1 (I) chain (*P02452*) |
| **e10563** | COL2A1 | NPGEPGEpGVSGPMGpRGPpGP | collagen alpha-1 (II) chain (*P02458*) |
| **e11829** | COL2A1 | GETGAAGpPGpAGPAGERGEQGAPGP | collagen alpha-1 (II) chain (*P02458*) |
| **e03506** | COL3A1 | SpGERGETGppGP | collagen alpha-1 (III) chain (*P02461*) |
| **e05700** | COL3A1 | TGpGGDKGDTGPpGPQG | collagen alpha-1 (III) chain (*P02461*) |
| **e07668** | COL3A1 | GTGGPpGENGKpGEpGPKG | collagen alpha-1 (III) chain (*P02461*) |
| **e11222** | COL3A1 | NDGApGKNGERGGpGGpGPQGPpG | collagen alpha-1 (III) chain (*P02461*) |
| **e18839** | COL3A1 | GPPGMPGPRGSPGpQGVKGESGKpGANGLSGERGpPGPQG | collagen alpha-1 (III) chain (*P02461*) |
| **e05473** | COL4A1 | GPpGFTGPPGPPGPPGP | collagen alpha-1 (IV) chain (*P02462*) |
| **e02022** | COL5A1 | KGNSGGDGpAGPP | collagen alpha-1 (V) chain (*P20908*) |

**Table S2  Urinary Peptide Sequences Retained in the Analyses (Ends)**

| ID | Symbol | Amino-Acid Sequence | Protein (*UniProt ID*) |
|---|---|---|---|
| **e16281** | COL6A1 | GADGEAGRPGSSGPSGDEGQPGEpGppGEKGEA | collagen alpha-1 (VI) chain (*P12109*) |
| **e17131** | COL6A1 | PPGDPGLMGERGEDGpAGNGTEGFpGFPGYPGN | collagen alpha-1 (VI) chain (*P12109*) |
| **e01100** | COL7A1 | DRGEpGPpGP | collagen alpha-1 (VII) chain (*Q02388*) |
| **e12945** | COL11A1 | DGpQGPpGSVGSVGGVGEKGEPGEAGN | collagen alpha-1 (XI) chain (*P12107*) |
| **e02311** | COL15A1 | GpPGPpGPpGPpA | collagen alpha-1(XV) chain (*P39059*) |
| **e05421** | COL16A1 | HpGppGEPGTDGAAGK | collagen alpha-1 (XVI) chain (*Q07092*) |
| **e18049** | COL22A1 | RGESGAMGLPGQEGLPGKDGDTGPTGPQGpQGpRGp | collagen alpha-1 (XXII) chain (*Q8NFW1*) |
| **e02933** | COL1A2 | GppGPDGNKGEpG | collagen alpha-2 (I) chain (*P08123*) |
| **e04987** | COL1A2 | pGpQGVQGGKGEQGp | collagen alpha-2 (I) chain (*P08123*) |
| **e10651** | COL1A2 | PpGKAGEDGHpGKPGRpGERG | collagen alpha-2 (I) chain (*P08123*) |
| **e09063** | COL5A2 | GARGLTGNpGVQGpEGKLGP | collagen alpha-2 (V) chain (*P05997*) |
| **e16610** | COL4A2 | DTGNPGAPGTpGTKGWAGDSGpQGRpGVFGLPG | collagen alpha-2 (IV) chain (*P08572*) |
| **e09267** | COL5A2 | PGPVGApGDAGQRGDPGSRGP | collagen alpha-2 (V) chain (*P05997*) |
| **e20509** | COL11A2 | GEHGpPGPPGPIGPVGQPGAAGADGEPGARGPQGHFGAKGDEGTRGFNGP | collagen alpha-2 (XI) chain (*P13942*) |
| **e15360** | COL4A3 | GpKGDpGIpGLDRSGFpGETGSPGIPGHQ | collagen alpha-3 (IV) chain (*Q01955*) |
| **e16874** | COL5A3 | DLGPpGDpGVSGIDGSpGEKGDPGDVGGPGPPGASG | collagen alpha-3 (V) chain (*P25940*) |
| **e02189** | COL4A4 | GpPGPpGPpGPpG | collagen alpha-4 (IV) chain (*P12109*) |
| **e06373** | COL4A6 | SGpPGFPGLGTTGEKGE | Collagen alpha-6(IV) chain (*Q14031*) |

A lower case “p” in the amino-acid sequence indicates hydroxyproline.  The protein identification number was obtained from the UniProt database (www.uniprot.org).

**Table S3 Comparison of the Baseline Urinary Peptide Levels between Randomized Groups (Starts)**

| IDs of Selected Peptides | HOMAGE-RCT | | | PRIORITY-RCT | | |
|---|---|---|---|---|---|---|
| | **Placebo** | **Spironolactone** | ***P*** | **Placebo** | **Spironolactone** | ***P*** |
| **N° Patients** | 144 | 146 | … | 61 | 49 | … |
| **Selected in HOMAGE-RCT** | | | | | | |
| **e04960** | 4.81 (4.31-33.9) | 4.81 (4.71-37.8) | 0.8065 | 5.27 (4.76-8.08) | 5.26 (5.17-5.82) | 0.9514 |
| **e08916** | 2133 (1644-2984) | 1981 (1495-2649) | 0.1958 | 1670 (1169-2551) | 1640 (1134-2365) | 0.2366 |
| **e09408** | 725 (415-1297) | 679 (455-1034) | 0.3772 | 818 (453-1271) | 910 (568-1262) | 0.3322 |
| **e10266** | 14,824 (10,672-20,817) | 15,567 (9956-20,058) | 0.4032 | 16,364 (13,628-21,661) | 15,737 (11,323-21,779) | 0.5588 |
| **e10395** | 6150 (4872-7577) | 5887 (4909-7096) | 0.2870 | 7330 (4781-9225) | 6510 (5377-9429) | 0.7846 |
| **e10437** | 415 (197-649) | 374 (170-637) | 0.8641 | 250 (86.8-392) | 277 (77.0-434) | 0.8110 |
| **e10657** | 691 (309-1245) | 488 (211-1084) | 0.2580 | 277 (84.9-698) | 275 (116-717) | 0.8091 |
| **e10863** | 2901 (2127-3916) | 2803 (2071-3953) | 0.9614 | 2313 (1485-3301) | 2192 (1286-3066) | 0.5469 |
| **e11972** | 3179 (1469-4696) | 3267 (1818-4775) | 0.7468 | 2206 (1209-2663) | 1403 (830-2544) | 0.4194 |
| **e17425** | 45.4 (11.0-126) | 55.9 (11.1-151) | 0.4181 | 29.9 (29.8-95.3) | 29.9 (29.0-79.7) | 0.3934 |
| **e18740** | 6.50 (6.49-47.9) | 6.62 (6.50-43.2) | 0.4241 | 9.67 (9.51-39.9) | 9.67 (8.49-42.9) | 0.8901 |
| **e10563** | 129 (8.15-312) | 109 (8.15-312) | 0.8203 | 3.78 (3.78-118.6) | 87.2 (3.78-417) | 0.0662 |
| **e03506** | 190 (63.5-351) | 196 (66.5-355) | 0.6131 | 141.8 (4.44-236.8) | 231 (104-327) | 0.0416 |
| **e05700** | 5.17 (4.36-327) | 5.52 (5.36-305) | 0.8772 | 20.0 (19.8-93.1) | 20.1 (19.6-68.6) | 0.4766 |
| **e07668** | 60.5 (13.3-274) | 17.7 (13.3-259) | 0.8744 | 45.7 (6.89-327) | 6.89 (6.89-172) | 0.0719 |
| **e18839** | 209 (23.8-519) | 193 (63.9-486) | 0.3347 | 219 (32.6-805) | 157 (6.22-409) | 0.2853 |
| **e05473** | 2.62 (2.44-26.7) | 2.62 (2.44-50.5) | 0.1945 | 3.78 (3.40-3.78) | 3.78 (3.40-17.9) | 0.3430 |

**Table S3 Comparison of the Baseline Urinary Peptide Levels between Randomized Groups (Continued)**

| Peptide IDs | HOMAGE-RCT | | | PRIORITY-RCT | | |
|---|---|---|---|---|---|---|
| | **Placebo** | **Spironolactone** | ***P*** | **Placebo** | **Spironolactone** | ***P*** |
| **e17131** | 156 (61.1-347) | 157.1 (3.10-339.1) | 0.5213 | 53.1 (6.06-143) | 63.2 (6.06-168) | 0.7980 |
| **e01100** | 94.7 (4.85-253) | 87.7 (4.85-206.3) | 0.3725 | 22.6 (3.96-136) | 49.2 (3.96-142) | 0.6653 |
| **e02933** | 468 (273-619) | 441 (295-656) | 0.7174 | 560 (360-673) | 581 (394-835) | 0.2320 |
| **e04987** | 66.6 (9.97-279) | 43.9 (9.97-290.4) | 0.7897 | 15.9 (15.6-136) | 15.9 (15.4-107) | 0.4451 |
| **e16610** | 5.26 (4.89-23.5) | 5.26 (4.88-31.7) | 0.5890 | 6.60 (5.87-6.68) | 6.68 (6.57-6.69) | 0.4010 |
| **e09267** | 3.34 (2.96-10.8) | 3.34 (3.08-13.3) | 0.9044 | 6.90 (6.62-11.4) | 6.90 (6.25-7.34) | 0.3652 |
| **e20509** | 879 (362-1776) | 1025 (444-1858) | 0.7942 | 1875 (555-3927) | 1981 (640-4464) | 0.5935 |
| **e15360** | 358 (175-678) | 339 (176-643) | 0.6530 | 222 (73.2-426) | 194 (54.5-421) | 0.9603 |
| **e16874** | 4.56 (4.07-17.0) | 4.56 (3.75-20.8) | 0.5382 | 3.62 (2.86-4.52) | 3.64 (3.02-6.64) | 0.1678 |
| **e06373** | 190 (72.8-367) | 230 (88.0-409) | 0.3494 | 212 (151-332) | 176 (59.3-396) | 0.0552 |
| **Selected in PRIORITY-RCT** | | | | | | |
| **e10142** | 278 (104-428) | 269 (125-398) | 0.4654 | 216 (73.8-407) | 192 (88.7-321) | 0.6400 |
| **e11829** | 6.35 (6.07-56.7) | 6.30 (5.70-23.7) | 0.0382 | 9.80 (9.19-9.81) | 9.80 (9.23-14.3) | 0.3990 |
| **e11222** | 303 (134-431) | 321 (65.3-554) | 0.5256 | 177.2 (72.1-369.6) | 171 (46.2-280) | 0.3076 |
| **e02022** | 3.97 (3.19-28.7) | 3.97 (3.34-25.9) | 0.5584 | 9.69 (8.89-10.5) | 9.59 (8.99-10.5) | 0.9468 |
| **e16281** | 235 (83.6-460) | 192 (64.4-518) | 0.8343 | 86.7 (27.3-220) | 127 (27.3-250) | 0.3907 |
| **e12945** | 4.46 (3.85-86.2) | 4.42 (3.89-33.3) | 0.0207 | 2.38 (1.62-2.38) | 2.38 (1.99-50.6) | 0.2174 |
| **e02311** | 1418 (616-2363) | 1380 (679-2800) | 0.4306 | 562 (134-1036) | 619 (47.3-1242) | 0.8817 |
| **e05421** | 109 (10.1-221) | 96.8 (12.9-239) | 0.9710 | 4.19 (4.13-32.2) | 4.22 (3.86-12.6) | 0.2968 |
| **e18049** | 406 (171-2576) | 427 (171-2759) | 0.9071 | 38.0 (37.2-1078) | 38.0 (37.2-941) | 0.9382 |

**Table S3 Comparison of the Baseline Urinary Peptide Levels between Randomized Groups (Ends)**

| Peptide IDs | HOMAGE-RCT | | | PRIORITY-RCT | | |
|---|---|---|---|---|---|---|
| | Placebo | Spironolactone | *P* | Placebo | Spironolactone | *P* |
| **e10651** | 36.0 (5.37-124.7) | 40.7 (5.37-134) | 0.9633 | 43.8 (4.40-172) | 17.2 (4.40-138) | 0.3533 |
| **e09063** | 44.3 (15.7-227) | 15.7 (15.7-208) | 0.3559 | 222 (19.2-678) | 241 (19.2-524) | 0.7914 |
| **e02189** | 458 (95.5-631) | 354 (128-811) | 0.2407 | 205 (119-431) | 89.9 (3.91-445) | 0.0492 |
| **e06373** | 190 (72.8-368) | 230 (88.0-409.3) | 0.3494 | 212 (151-332) | 176 (59.3-396) | 0.0552 |

Tabulated values are median (interquartile range) of the urinary proteomic markers retained in the statistical analyses of HOMAGE-RCT (Table 3) and PRIORITY-RCT (Table 4). Peptide e06373 was retained in both trials. *P*-values were computed by the large sample Z test based on rank-normalized peptide levels.

**Table S4 Correlations Between the Changes over Follow-up in COL1A1-Derived Urinary Peptides and in CITP in the HOMAGE Subset**

| Biomarkers | Placebo (n=144) | | Spironolactone (n=146) | | $P_{DIF}$ |
|---|---|---|---|---|---|
| | ***r*** **(95% CI)** | ***P*** | ***r*** **(95% CI)** | ***P*** | |
| **Increasing Peptide Levels** | | | | | |
| Δ **e08916** (*COL1A1*) | 0.126 (-0.039, 0.284) | 0.1319 | 0.256 (0.097, 0.402) | 0.0017 | 0.2548 |
| Δ **e09408** (*COL1A1*) | 0.146 (-0.018, 0.302) | 0.0799 | 0.286 (0.130, 0.429) | 0.0004 | 0.2150 |
| Δ **e10266** (*COL1A1*) | 0.104 (-0.061, 0.263) | 0.2151 | 0.168 (0.006, 0.322) | 0.0418 | 0.2826 |
| Δ **e10395** (*COL1A1*) | 0.147 (-0.017, 0.303) | 0.0782 | 0.101 (-0.062, 0.259) | 0.2234 | 0.6938 |
| Δ **e10437** (*COL1A1*) | 0.214 (0.053, 0.365) | 0.0095 | 0.122 (-0.042, 0.279) | 0.1422 | 0.4247 |
| Δ **e10657** (*COL1A1*) | 0.149 (-0.015, 0.305) | 0.0730 | 0.309 (0.154, 0.449) | 0.0001 | 0.1537 |
| Δ **e10863** (*COL1A1*) | 0.217 (0.055, 0.367) | 0.0086 | 0.297 (0.141,0.438) | 0.0002 | 0.4701 |
| **Decreasing peptide levels** | | | | | |
| Δ **e04960** (*COL1A1*) | -0.021 (-0.184, 0.143) | 0.8010 | 0.032 (-0.131, 0.193) | 0.7024 | 0.6551 |
| Δ **e11972** (*COL1A1*) | -0.057 (-0.219, 0.108) | 0.4968 | -0.046 (-0.207, 0.117) | 0.5810 | 0.9260 |
| Δ **e17425** (*COL1A1*) | -0.070 (-0.231, 0.095) | 0.4047 | -0.085 (-0.244, 0.079) | 0.3066 | 0.8988 |
| Δ **e18740** (*COL1A1*) | -0.147 (-0.303, 0.017) | 0.0779 | 0.049 (-0.114, 0.210) | 0.5523 | 0.0967 |

Abbreviations: Δ, change from baseline to Month 9; CITP, collagen type I C-terminal telopeptide. Tabulated values are Pearson correlation coefficients (*r*), given with 95% confidence interval. Peptide levels increasing or decreasing from baseline to Month 9 are listed in Table 3. *P* and $P_{DIF}$ refer to the significance of the correlation coefficients and the significance of the between-group differences in the correlation coefficients, obtained by Fisher transformation.

**Table S5 Between-Group Differences in Serum Electrolytes and Renal Function at Last Follow-Up Visit**

| Marker | Placebo (n=144) | | Spironolactone (n=146) | | Δ (95% CI) | *P*-Value |
|---|---|---|---|---|---|---|
| | Median (IQR) | LMean±SE | Median (IQR) | LMean±SE | | |
| **HOMAGE** | | | | | | |
| Serum $Na^+$, mmol/L | --- | 139.3±0.2 | --- | 138.4±0.2 | -0.904 (-1.364, -0.443) | 0.0001 |
| Serum $K^+$, mmol/L | --- | 4.39±0.03 | --- | 4.53±0.03 | 0.139 (0.058, 0.219) | 0.0008 |
| eGFR, mL/min/1.73 $m^2$ | --- | 70.67±0.86 | --- | 68.18±0.85 | -2.49 (-4.94, -0.47) | 0.0468 |
| **PRIORITY** | | | | | | |
| Serum $Na^+$, mmol/L | --- | 140.3±0.4 | --- | 139.8±0.4 | -0.531 (-0.622, +1.682) | 0.3690 |
| Serum $K^+$, mmol/L | --- | 4.25±0.05 | --- | 4.44±0.06 | +0.182 (+0.028, +0.337) | 0.0229 |
| eGFR, mL/min/1.73 $m^2$ | --- | 78.6±1.3 | --- | 75.2±1.5 | -3.22 (-7.38, +0.93) | 0.1315 |
| UACR, mg/g | 7.64 (4.57-15.87) | 0.017±0.127 | 7.24 (4.38-14.83) | -0.021±0.144 | -0.038 (-0.437, +0.360) s | 0.8510 |

Abbreviations: eGFR, glomerular filtration rate estimated from serum creatinine according to the Chronic Kidney Disease Epidemiology equation; IQR, interquartile range; LMean, least square mean; SE, standard error; UACR, urine albumin-creatinine ratio; Δ (95% CI), between-group difference (spironolactone minus placebo) given with 95% confidence interval; *P*-value, significance of the between-group differences. Median (IQR) UACR was derived from the "*real world*" non-normalized non-adjusted data. LMean±SE and Δ (95% CI) rest on non-normalized or rank normalized data for UACR. Δ (95% CI) provides multivariable-adjusted estimates of the between-group differences in "*real world*" units or standardized units for UACR. All models were adjusted for sex, age, BMI, and the baseline value of the marker. The other covariables in HOMAGE analyses included current smoking and drinking, history of ischemic heart disease, and treatment at baseline and change in treatment at the last follow-up with antihypertensive, lipid-lowering, antiplatelet and antidiabetic drugs. The other covariables in PRIORITY analyses included smoking, history of cardiovascular disease, and treatment at baseline with antihypertensive and antidiabetic drugs.

234
